# Analysis of severe outcomes associated with the SARS-CoV-2 Variant of Concern 202012/01 in England using ICNARC Case Mix Programme and QResearch databases

**DOI:** 10.1101/2021.03.11.21253364

**Authors:** Martina Patone, Karen Thomas, Rob Hatch, Pui San Tan, Carol Coupland, Weiqi Liao, Paul Mouncey, David Harrison, Kathryn Rowan, Peter Horby, Peter Watkinson, Julia Hippisley-Cox

**Author notes:** joint senior authors (Peter Watkinson, Julia Hippisley-Cox).

## Abstract

**Background:** A new, more transmissible variant of SARS-CoV-2, variant of concern (VOC) 202012/01 or lineage B.1.1.7, has emerged in the UK. We estimate the risk of critical care admission, mortality in critical ill patients, and overall mortality associated with VOC B.1.1.7 compared with the original variant. We also compare clinical outcomes between these variants ‘ groups.

**Methods:** We linked a large primary care (QResearch), the national critical care (ICNARC CMP) and the COVID-19 testing (PHE) database and extracted two cohorts. The first was used to explore the association between VOC B.1.1.7 and critical care admission and 28-day mortality. The second to determine the risk of mortality in critically ill patients with VOC B.1.1.7 compared to those without. We used Royston-Parmar models adjusted for age, sex, region, other socio-demographics and comorbidities (asthma, COPD, type I and II, hypertension). We reported information on types and duration of organ supports for the two variants ‘ groups.

**Findings:** The first cohort included 198,420 patients. Of these, 80,494 had VOC B.1.1.7, 712 were critically ill and 630 died by 28 days. The second cohort included 3432 critically ill patients. Of these, 2019 had VOC B.1.1.7 and 822 died at the end of critical care. Using the first cohort, we estimated adjusted hazard ratios for critical care admission and mortality to be 1.99 (95% CI: 1.59, 2.49) and 1.59 (95% CI: 1.25-2.03) for VOC B.1.1.7 compared with the original variant group, respectively. The adjusted hazard ratio for mortality in critical care, estimated using the second cohort, was 0.93 (95% CI 0.76-1.15) for patients with VOC B.1.1.7, compared to those without.

**Interpretation:** VOC B.1.1.7 appears to be more severe. Patients with VOC B.1.1.7 are at increased risk of critical care admission and mortality compared with patients without. For patients receiving critical care, mortality appears independent of virus strain.

**RESEARCH IN CONTEXT:** *Evidence before this study:* A new variant of the SARS-CoV-2 virus, variant of concern (VOC) 202012/01, or lineage B.1.1.7, was detected in England in September 2020. The characteristics and outcomes of patients infected with VOC B.1.1.7 are not yet known. VOC B.1.1.7 has been associated with increased transmissibility. Early analyses have suggested infection with VOC B.1.1.7 may be associated with a higher risk of mortality compared with infection with other virus variants, but these analyses had either limited ability to adjust for key confounding variables or did not consider critical care admission. The effects of VOC B.1.1.7 on severe COVID-19 outcomes remain unclear.

*Added value of this study:* This study found a 60% higher risk of 28-day mortality associated with infection with VOC B.1.1.7 in patients tested in the community in comparison with the original variant, when adjusted for key confounding variables. The risk of critical care admission for those with VOC B.1.1.7 is double the risk associated with the original variant. For patients receiving critical care, the infecting variant is not associated with the risk of mortality at the end of critical care.

*Implications of all the available evidence:* The higher mortality and rate of critical care admission associated with VOC B.1.1.7, combined with its known increased transmissibility, are likely to put health care systems under further stress. These effects may be mitigated by the ongoing vaccination programme.

## BACKGROUND

Infection with the SARS-CoV-2 virus has resulted in large numbers of patients with COVID-19 receiving critical care. More than 34,000 such patients have received critical care in England^1^. Worldwide, mortality following critical illness with COVID-19, up to May 2020, was reported as 40-50%^2^. A more recent systematic review^3^ indicated that mortality following critical illness with COVID-19 was lower at the end of September 2020 (35%) than at the end of May 2020 (42%).

In September 2020, a new variant of the SARS-CoV-2 virus, known as variant of concern (VOC) 202012/01, or lineage B.1.1.7, was detected by the COVID-19 Genomics UK (COG-UK) consortium in England^4^. VOC B.1.1.7 has multiple changes, including an N501Y substitution in the spike protein that enhances binding to the human ACE2 receptor, which the virus uses to enter the cell. It has been suggested that these changes may result in increased infectivity^5^, with initial reports of 50-74% increased transmissibility^6^. The characteristics and outcomes of patients infected with VOC B.1.1.7 are not yet known. Early analyses, of mortality linked to diagnostic data, have suggested that infection with VOC B.1.1.7 may be associated with a higher risk of mortality compared with infection with other virus variants^7,8,9,10,11,12^. However, these analyses had either limited adjustment for key patient characteristics thought to be associated with COVID-19 outcomes, or did not consider effects on critical care admission or outcome. The effect of VOC B.1.1.7 on severe COVID-19 outcomes, receipt of critically care and mortality, carefully adjusted for key patient characteristics, remains unclear. VOC B.1.1.7 has now been detected worldwide^13^.

We explored the association between VOC B.1.1.7 and the risk of receiving critical care and 28-day mortality, following a positive community COVID-19 test. In addition, for patients with confirmed COVID-19 receiving critical care, we explored the association between VOC B.1.1.7 and receipt and duration of organ support in critical care, duration of critical care stay and mortality at the end of critical care. This analysis is part of a larger study protocol ^14^.

## METHODS

### Data platform

The QResearch data platform is a high-quality, research database based on records from 1350 primary care practices in England. Established in 2002, QResearch has been used extensively for epidemiological research^15^. QResearch is one of the largest, and most representative, primary care research databases, nationally^16^ covering approximately 20% of the population of England. It has been used for COVID-19 research to inform the national pandemic response in the first pandemic wave^17, 18^.

### Data linkage

To ensure that the QResearch data platform could be used to inform policy and planning during the UK COVID-19 epidemic, the primary care data were linked to other databases. The key data linkages for this research were: (1) COVID-19 testing data (the national registry of COVID-19 RT-PCR positive test results from Public Health England (PHE)) - COVID-19 is a notifiable disease and laboratories in England are required to send results of all tests to PHE; (2) the ICNARC COVID-19 study data consisting of critically ill patients with confirmed COVID-19 (confirmed at/or on admission to critical care) hosted on the ICNARC Case Mix Programme (the national, high-quality clinical database for adult critical care) with complete coverage of critical care units across England (Wales and Northern Ireland); and (3) Office of National Statistics COVID-19 mortality data, which includes all deaths due to COVID-19 in England.

### VOC B.1.1.7

As part of test results sent to PHE, the molecular diagnosis of SARS-CoV-2 used is included. VOC B.1.1.7 has a deletion of six nucleotides in the S-gene that results in the deletion of two amino acids at positions 69 and 70 of the spike glycoprotein, which leads to S-gene molecular diagnostic assay failure (SGTF).

Cycle threshold (Ct) values for the S, N, and ORF1ab components of SARS-CoV-2, used to define the SGTF status, are available for COVID-19 RT-PCR positive test taken in the community (not hospital) setting. We define SGTF as any test with non-detectable S gene and Ct <= 30 for N and ORF1ab target and non-SGTF as any test with detectable S gene and Ct <= 30 for N and ORF1ab target. All other tests were defined as inconclusive and excluded from the analysis. Currently, in the UK, greater than 99% of SGTF are due to VOC B.1.1.7^8^. PCR positive samples with SGTF, therefore, were used as a proxy to identify the presence (or absence) of VOC B.1.1.7.

### Patient cohorts

Our full study observation period was 1 November 2020 to 27 January 2021. We selected a start date based on the emergence of VOC B.1.1.7 i.e. 99% of patients with VOC B.1.1.7 were identified after this date (Supplementary Figure 1). Data on critically ill admissions were available up to 27 January 2021 and ONS mortality data up to 31^st^ January 2021. We chose to censor mortality data five days before the last data update to reduce the effect of late reporting bias.

We extracted two cohorts of patients from the linked data to explore the association of VOC B.1.1.7 with severe COVID-19 outcomes:

- The ‘primary care cohort’ was patients in primary care with a positive community COVID-19 test reported between 1 November 2020 and 26 January 2021.
- The ‘critical care cohort’ was patients admitted for critical care with a positive community COVID-19 test reported between 1 November 2020 and 27 January 2021.

We used the primary care cohort to determine the association of VOC B.1.1.7 with receipt of critical care and with the risk of 28-day mortality. We used the critical care cohort to determine the association of VOC B.1.1.7 with duration of organ support in critical care, duration of critical care and mortality at the end of critical care.

The patient cohorts are reported according to the RECORD guidelines ^19^.

### Data

Key patient characteristics, for which data were extracted, for the primary care cohort were:

- age (years);
- sex (male, female);
- ethnic group (White, Indian, Pakistani, Bangladeshi, Other Asian, Caribbean, Black African, Chinese, Other ethnic groups);
- body mass index (BMI grouped: <25, 25-30, 30-40, >=40);
- co-morbidities (asthma, COPD, diabetes type I and II, hypertension);
- smoking status (non, ex-, light, moderate and heavy);
- deprivation (quintile, based on Townsend score);
- housing category (care home, homeless or neither);
- household size (1, 2, 3-5, 6+ people); and
- geographical region (10 across England).

The outcomes of interest for the primary care cohort were receipt of critical care and 28-day mortality with COVID-19 defined as confirmed or suspected COVID-19 recorded on the death certificate or death from any cause within 28 days of a positive COVID-19 test.

Key patient characteristics, for which data were extracted, for the critical care cohort were:

- age (years);
- sex (male, female);
- ethnic group (White, Indian, Pakistani, Bangladeshi, Other Asian, Caribbean, Black African, Chinese, Other ethnic groups);
- co-morbidities (cardiovascular, respiratory, metastatic disease, immunocompromised);
- dependency (assistance with activities of daily living – none, some, all);
- pregnancy (currently pregnant, recently pregnant in last six weeks, not known to be pregnant); and
- cardiopulmonary resuscitation (in 24 hours prior to critical care admission).

The outcomes of interest for the critical care cohort were duration of organ support (respiratory, cardiovascular, renal, neurological and liver) in critical care, duration of critical care and mortality at the end of critical care.

## STATISTICAL ANALYSIS

### Royston-Parmar models

We used flexible parametric survival models (Royston-Parmar model) to estimate the hazard ratio (HR) for 28-days mortality and admission to critical care comparing patients with VOC B.1.1.7 and without in the primary-care cohort, and for mortality at the end of critical care in the critical care cohort.

In all the models, degrees of freedom were chosen to minimise the Akaike information criterion (AIC) and possible interactions between VOC B.1.1.7 and age, sex and ethnicity were tested using the Wald test. When the proportional hazard assumption was not met, a time varying hazard ratio was modelled.

For each cohort, we accounted for missing data by using multiple imputation by chained equations, which generated five imputed datasets each. The imputation model included age, sex, the outcome of interest (28-day mortality, critical care admission or critical care mortality), and all confounding and mediating variables. We fitted Royston-Parmar models within each imputed dataset and combined them in accordance with Rubin ‘s rules.

A post-hoc power calculation shows there is 80% power at the 0.05 significance level to detect a hazard ratio for admission to critical care in the VOC B.1.1.7 group of > 1.04 or less than 0.96, and for mortality at the end of critical care in the VOC B.1.1.7 group of > 1.09 or less than 0.91.

### Adjustments

For estimating the HR of 28-day mortality and critical care admission in the primary-care cohort, the models were adjusted for patients ‘ demographics (age, sex, deprivation index, geographical region, ethnicity, house size, BMI and smoking status) and co-morbidities (asthma, COPD, diabetes type I and II, hypertension). Age was modelled using a restricted cubic spline. To account for time dependent biases, positive test date was included in the models. A random effect frailty term was included to account for similarities amongst patients registered in the same GP practice.

The model used to estimate the association between VOC B.1.1.7 and risk of mortality at the end of critical care was adjusted by age, sex, ethnicity, BMI, severe comorbidities, dependency prior to admission to acute hospital and geographic region. The date of admission to critical care was included in the model to account for time dependent biases. A random frailty term was included to account for similarities amongst patients registered in the same critical care unit. Including the age variable as a linear term rather than a restricted cubic spline did not change the results and hence the linear term is included in the final model.

### Censoring

The start time for the mortality and admission to critical care analyses using the primary care cohort was the date of positive test. Patients were followed for 20 days when investigating the relative risk of admission for critical care, and 28 days when exploring the relative risk of mortality between the two variants’ groups. Individuals who did not receive critical care within 20 days (for critical care admission analysis) or who did not die within 28 days (for mortality analysis) were censored at 20, or 28 days respectively, after the date of positive test. Patients who died during the follow-up period before receiving critical care or for non-COVID-19 causes were censored at their date of death.

For the analysis of mortality at the end of critical care in the critical care cohort, the start date was the date of their critical care admission. Patients were censored after 28 days and those who survived were censored on their date of discharge from critical care.

### Sensitivity analyses

Patients with VOC B.1.1.7 became the majority at the end of the study period (Supplementary Figure 1).This caused patients with VOC B.1.1.7 to have a shorter follow-up time and possibly to have their outcome not completed by the end of the study. To assure that the outcome was known for all patients, with and without VOC B.1.1.7, we run a sensitive analysis for each outcome. In the primary care analysis, we restricted the cohort to only patients who had completed their follow-up of 20 and 28 days for the critical care admission and mortality analysis, respectively. In the critical care analysis, we included only patients who had completed their critical care outcomes (death or survival at discharge).

All analyses were re-run including only the complete case dataset as additional sensitivity analysis.

### A matched cohort for reporting critical care outcomes

The outcome of 28-day mortality was chosen as it was available for all patients irrespective of date of admission, in contrast to information on types and duration of organ support for which there was more availability in patients who were admitted earlier on in the study period. Hence, to compare the clinical characteristics of patients in the VOC B.1.1.7 group and in the non-VOC B.1.1.7 group we restricted our reporting to a matched cohort of patients, derived from the critical care cohort.

Each critically ill patient with VOC B.1.1.7 was matched with a critically ill patient without VOC B.1.1.7 admitted to the same unit. Only pair of patients who were admitted within 3 days of each other were kept. If a patient with VOC B.1.1.7 was matched with more than one patient without VOC B.1.1.7, one pair only was randomly selected.

## RESULTS

### Primary care cohort

During the study period (1 November 2020 to 26 January 2021), there were 12,278,186 patients registered with participating primary care practices in QResearch, 2,091,828 positive COVID-19 RT-PCR tests from PHE and 13,907 admissions for critical care in the ICNARC COVID-19 study. Combined, this produced a linked dataset of 429,926 patients, of which, 381,887 had a positive COVID-19 RT-PCR test in the community and SGTF status (as a proxy for VOC B.1.1.7) were identifiable in 198,420 (51.9 %) (Figure 1). Of these 198,420 patients, 613 patients had died by 28 days and 712 were admitted for critical care. The community test date was after the recorded date of death in only 13 patients.

**Figure 1:**
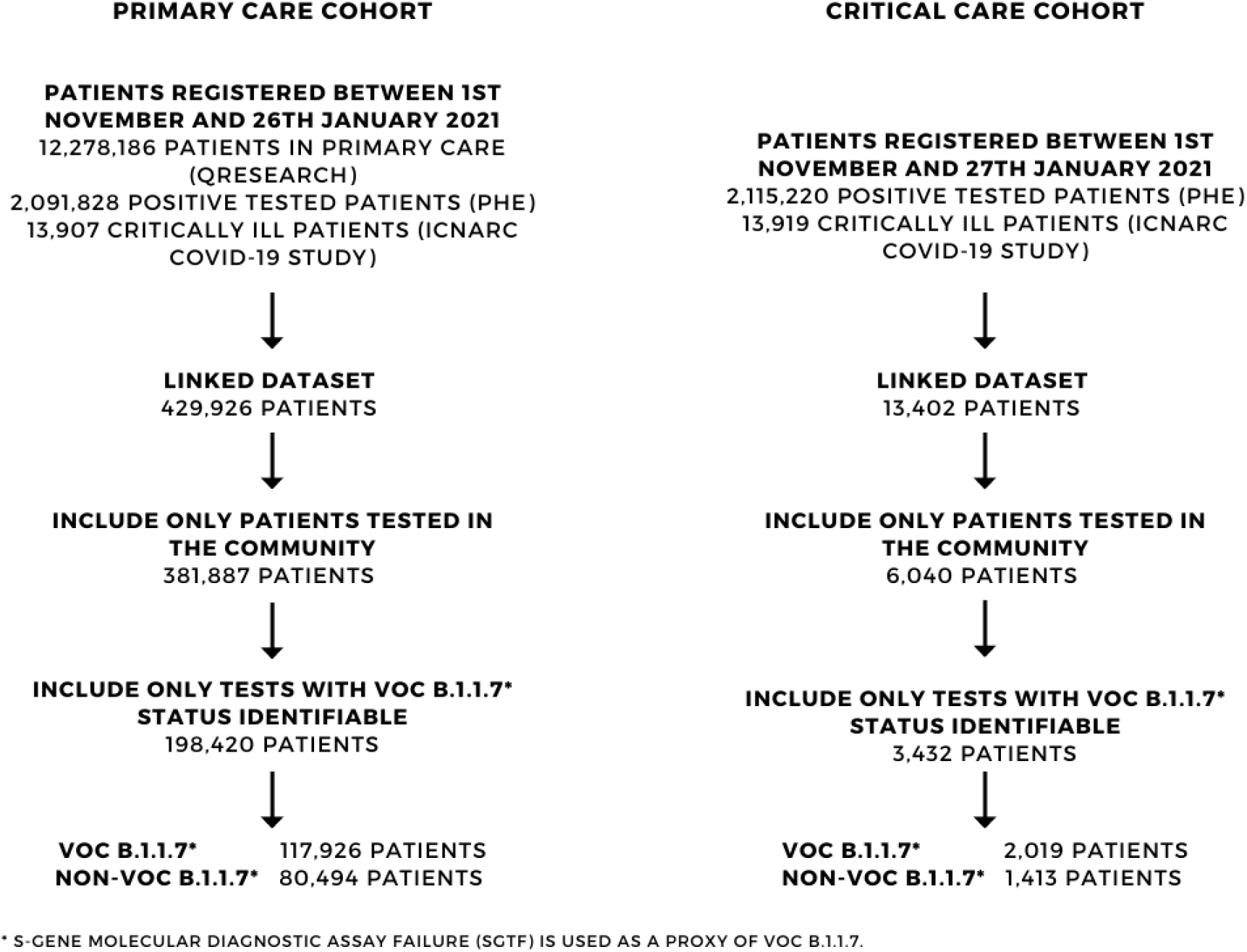
Flowchart describing inclusion in the primary care cohort and in the critical care cohort.

#### VOC B.1.1.7

Of the 198,420 patients for whom results were available, 117,926 (59.4%) had VOC B.1.1.7 and 80,494 had non-VOC B.1.1.7 (Figure 1).VOC B.1.1.7 became increasingly dominant over the study period (Supplementary Figure 1).

Table 1 presents the demographic and clinical characteristics for patients in the primary care cohort by VOC B.1.1.7. Patients in the VOC B.1.1.7 group and the non-VOC B.1.1.7 group were broadly similar, with some regional differences, however, the VOC B.1.1.7 group had a lower proportion of patients 70 years of age or above (3.6 % versus 4.7%).

**Table 1:** Demographics of primary care patients tested positive in the community between 1^st^ November 2020 to 26^th^ January 2021 (primary care cohor t), by variant.

|  | **Non-VOC B.1.1.7**<br>*Cols% (counts)* | **VOC B.1.1.7\***<br>*Cols% (counts)* | **Full Cohort**<br>*Cols% (counts)* |
| --- | --- | --- | --- |
| **Total number of patients** | 80494 | 117926 | 198420 |
| **ICU admitted** | 0.3 (263) | 0.4 (449) | 0.4 (712) |
| **Deaths within 28 days** | 0.3 (273) | 0.3 (340) | 0.3 (613) |
| **Sex** |  |  |  |
| Female | 53.4 (42976) | 52.3 (61679) | 52.7 (104655) |
| Male | 46.6 (37518) | 47.7 (56247) | 47.3 (93765) |
| **Mean age (SD)** | 38.0 (18.1) | 37.4 (17.6) | 37.7 (17.8) |
| **Age Categories** |  |  |  |
| 18-29 | 36.2 (29122) | 35.6 (42000) | 35.8 (71122) |
| 30-39 | 19.1 (15412) | 20.4 (24012) | 19.9 (39424) |
| 40-49 | 16.5 (13255) | 17.5 (20622) | 17.1 (33877) |
| 50-59 | 15.7 (12604) | 15.6 (18342) | 15.6 (30946) |
| 60-69 | 7.8 (6318) | 7.3 (8662) | 7.5 (14980) |
| 70-79 | 3.1 (2510) | 2.5 (2994) | 2.8 (5504) |
| 80-89 | 1.3 (1022) | 0.9 (1073) | 1.1 (2095) |
| 90-99 | 0.3 (251) | 0.2 (221) | 0.2 (472) |
| **Ethnicity** |  |  |  |
| White | 60.9 (49016) | 58.4 (68907) | 59.4 (117923) |
| Indian | 3.8 (3046) | 4.2 (4900) | 4.0 (7946) |
| Pakistani | 5.0 (4045) | 3.7 (4323) | 4.2 (8368) |
| Bangladeshi | 2.5 (2008) | 3.0 (3549) | 2.8 (5557) |
| Other Asian | 1.9 (1526) | 2.5 (2958) | 2.3 (4484) |
| Caribbean | 0.5 (400) | 1.1 (1339) | 0.9 (1739) |
| Black African | 1.6 (1295) | 2.4 (2803) | 2.1 (4098) |
| Chinese | 0.2 (159) | 0.3 (373) | 0.3 (532) |
| Other ethnic group | 3.3 (2647) | 4.4 (5162) | 3.9 (7809) |
| Not recorded | 20.3 (16352) | 20.0 (23612) | 20.1 (39964) |
| **Date of positive test** |  |  |  |
| 1 Nov to 14 Nov | 32.5 (26160) | 1.5 (1784) | 14.1 (27944) |
| 15 Nov to 28 Nov | 20.5 (16509) | 2.4 (2879) | 9.8 (19388) |
| 29 Nov to 12 Dec | 13.7 (10990) | 6.5 (7704) | 9.4 (18694) |
| 13 Dec to 26 Dec | 13.5 (10847) | 20.1 (23700) | 17.4 (34547) |
| 27 Dec to 10 Jan | 14.1 (11368) | 39.8 (46921) | 29.4 (58289) |
| 11 Jan to 26 Jan | 5.7 (4620) | 29.6 (34938) | 19.9 (39558) |
| **Household size** |  |  |  |
| 1 person | 26.8 (21580) | 25.8 (30432) | 26.2 (52012) |
| 2 people | 20.6 (16618) | 19.8 (23358) | 20.1 (39976) |
| 3-5 people | 45.0 (36192) | 46.8 (55186) | 46.1 (91378) |
| 6+ people | 7.6 (6104) | 7.6 (8950) | 7.6 (15054) |
| **House type** |  |  |  |
| Neither | 99.7 (80219) | 99.7 (117549) | 99.7 (197768) |
| Care home | 0.3 (226) | 0.2 (271) | 0.3 (497) |
| Homeless | 0.1 (49) | 0.1 (106) | 0.1 (155) |
| **BMI** |  |  |  |
| <25 | 54.4 (43759) | 55.0 (64816) | 54.7 (108575) |
| 25-30 | 13.2 (10654) | 12.5 (14706) | 12.8 (25360) |
| 30-40 | 5.6 (4528) | 5.1 (6007) | 5.3 (10535) |
| >= 40 | 3.2 (2540) | 2.7 (3151) | 2.9 (5691) |
| Not recorded | 23.6 (19013) | 24.8 (29246) | 24.3 (48259) |
| **Smoking status** |  |  |  |
| Non smoker | 57.4 (46182) | 55.5 (65447) | 56.3 (111629) |
| Ex smoker | 17.7 (14262) | 17.5 (20642) | 17.6 (34904) |
| Light smoker | 8.0 (6455) | 9.6 (11276) | 8.9 (17731) |
| Moderate smoker | 1.5 (1222) | 1.7 (2050) | 1.6 (3272) |
| Heavy smoker | 0.5 (429) | 0.6 (702) | 0.6 (1131) |
| Not recorded | 14.8 (11944) | 15.1 (17809) | 15.0 (29753) |
| **Geographical region in England** |  |  |  |
| East Midlands | 1.9 (1529) | 1.1 (1305) | 1.4 (2834) |
| East of England | 1.8 (1428) | 3.3 (3912) | 2.7 (5340) |
| London | 16.5 (13302) | 31.7 (37342) | 25.5 (50644) |
| North East | 5.1 (4066) | 1.9 (2194) | 3.2 (6260) |
| North West | 33.3 (26808) | 19.0 (22416) | 24.8 (49224) |
| South Central | 8.4 (6754) | 12.8 (15073) | 11.0 (21827) |
| South East | 4.5 (3609) | 15.2 (17945) | 10.9 (21554) |
| South West | 4.9 (3966) | 3.3 (3893) | 4.0 (7859) |
| West Midlands | 17.7 (14235) | 10.3 (12140) | 13.3 (26375) |
| Yorkshire & Humber | 6.0 (4797) | 1.4 (1706) | 3.3 (6503) |
| **Deprivation quintile** |  |  |  |
| 1 (least deprived) | 21.8 (17606) | 19.2 (22650) | 20.3 (40256) |
| 2 | 21.8 (17507) | 21.5 (25312) | 21.6 (42819) |
| 3 | 21.3 (17128) | 21.5 (25327) | 21.4 (42455) |
| 4 | 19.9 (16054) | 19.9 (23521) | 19.9 (39575) |
| 5 (most deprived) | 14.4 (11622) | 17.2 (20321) | 16.1 (31943) |
| Not recorded | 0.7 (577) | 0.7 (795) | 0.7 (1372) |
| **Comorbidities** |  |  |  |
| Asthma | 15.6 (12551) | 14.6 (17241) | 15.0 (29792) |
| COPD | 1.1 (875) | 0.8 (998) | 0.9 (1873) |
| Diabetes type 1 | 0.5 (440) | 0.5 (618) | 0.5 (1058) |
| Diabetes type 2 | 5.1 (4101) | 4.4 (5188) | 4.7 (9289) |
| Hypertension | 10.5 (8473) | 9.5 (11163) | 9.9 (19636) |
| Parkinson | 0.1 (64) | 0.1 (82) | 0.1 (146) |
| Epilepsy | 1.1 (872) | 1.1 (1278) | 1.1 (2150) |
| Cerebral palsy | 0.1 (73) | 0.1 (107) | 0.1 (180) |
| Motor Neuron Disease | <5 | 0.0 (5) | 0.0 (8) |
| Huntington's disease | <5 | 0.0 (5) | 0.0 (6) |
| Multiple sclerosis | 0.1 (115) | 0.1 (152) | 0.1 (267) |
| Myasthenia | 0.0 (20) | 0.0 (28) | 0.0 (48) |
| Downs syndrome | 0.0 (30) | 0.0 (51) | 0.0 (81) |
| Learning disabilities excluding Downs | 1.7 (1343) | 1.5 (1798) | 1.6 (3141) |

\*SGTF status was used as a proxy for VOC B.1.1.7.

#### Risk of COVID-19 28-day mortality

There were 340/117,926 (0.3%) deaths in the VOC B.1.1.7 group and 273/80494 (0.3%) in the non-VOC B.1.1.7 group. Supplementary Figure 1 shows the weekly deaths by VOC B.1.1.7 status over the study period. Figure 2 shows the Kaplan-Meier plot for risk of COVID-19 28-day mortality, by variant, for the complete case analysis.

**Figure 2:**
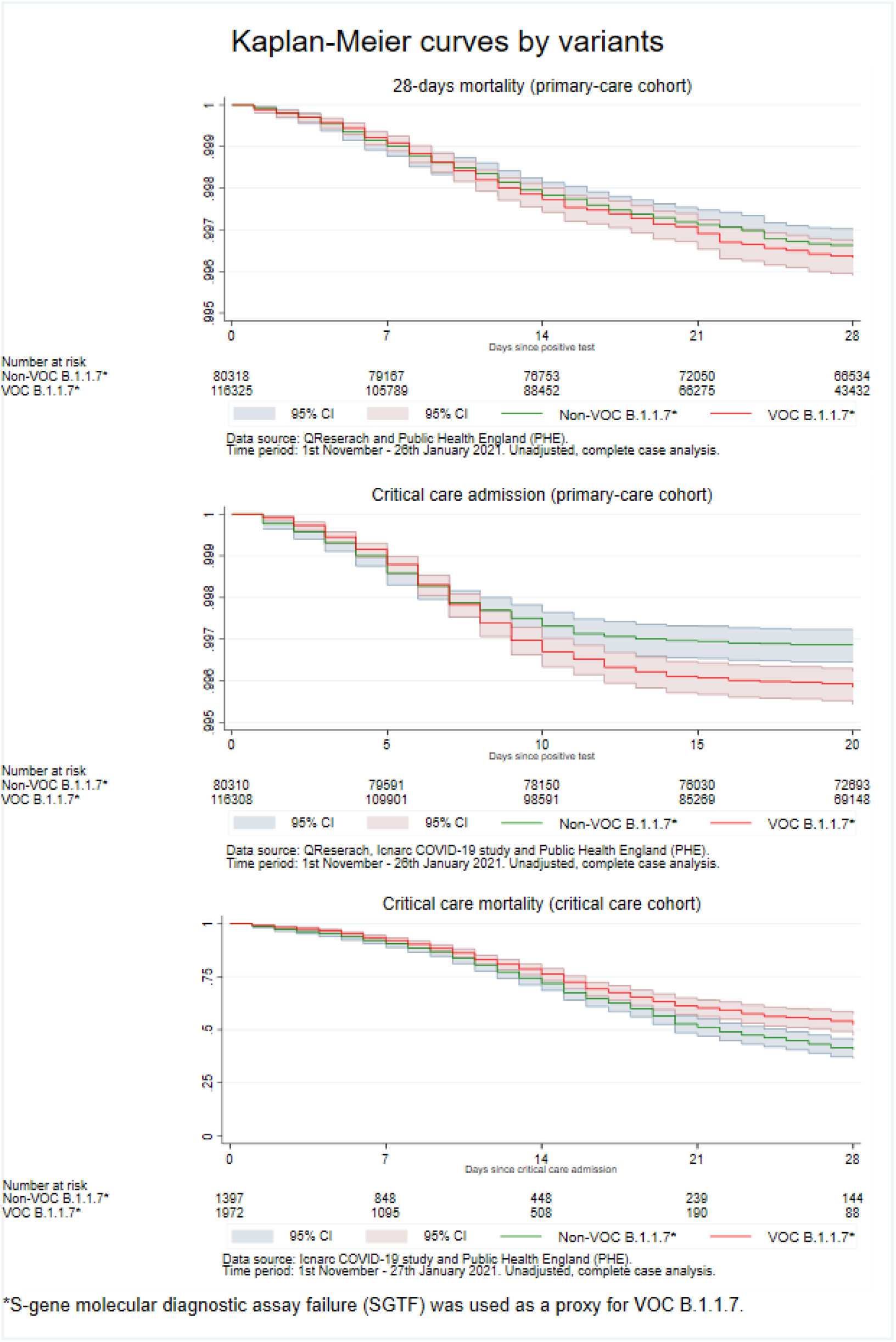
Kaplan-Meier plot for the risk of 28-days mortality, critical care admission for primary are patients tested positive in the community between 1^st^ November 2020 and 26^th^ January 2021 (primary care cohor t) and mortality at the end of critical care for critically ill patients tested positive in the community between 1^st^ November 2020 and 27^th^ January 2021 (critical care cohort). SGTF status is used as a proxy for VOC B.1.1.7.

Unadjusted analysis indicated no significant difference in COVID-19 28-day mortality risk for patients in the VOC B.1.1.7 (unadjusted HR: 1.09, 95% CI: 0.91-1.32) compared with those in the non-VOC B.1.1.7 group. After adjustment, we found a higher risk of COVID-19 28-day mortality (adjusted HR: 1.59; 95% CI: 1.25-2.03) for the VOC B.1.1.7 compared with the non-VOC B.1.1.7 group. The difference between the unadjusted and adjusted analyses was mainly explained by age (HR: 1.53, 95% CI: 1.28-1.82 with adjustment for age alone). We found no evidence of a significant interaction between VOC B.1.1.7 and: ethnic group (p = 0.39); sex (p = 0.71); or age group (p = 0.81). Adjusting only for the positive test date did not account for the increased risk of COVID-19 28-day mortality. Sensitivity analyses, including only those patients with at least 28-days follow-up, showed similar findings (adjusted HR: 1.64; 95% CI: 1.06-2.56) (the characteristics of the primary care cohort restricted to 28-days of follow-up are presented in supplementary Table S1).

#### Risk of critical care admission

In total, 712 patients were admitted for critical care in the primary care cohort. Of these, 449 (0.4%) were in the VOC B.1.1.7 group. Supplementary Figure 1 shows the weekly critical care admissions by VOC B.1.1.7 status over the study period. Figure 2 shows the Kaplan-Meier plot for risk of admission to critical care, by VOC B.1.1.7 group, for the complete case analysis.

Risk of admission to critical care was higher in the VOC B.1.1.7 compared with the non-VOC B.1.1.7 group in both unadjusted (HR: 1.27; 95% CI: 1.08 - 1.49) and adjusted (adjusted HR: 1.99; 95% CI: 1.59 - 2.49) analyses. However, the proportional hazard assumption was not met, so time varying HR was estimated and is presented in Figure 3. The time varying HR was 1.20 (95% CI: 0.58 - 2.48) one day after a positive test, 1.58 (95% CI: 1.13 - 2.21) five days after a positive test and 3.29 (95% CI: 1.17 - 6.29) and 2.80 (95% CI: 1.06 - 7.40) after fifteen and twenty days, respectively. We found no evidence of a significant interaction between VOC B.1.1.7 and: sex (p = 0.90), ethnic group (p = 0.64) or age group (p = 0.15). Adjusting only for the date of positive test did not account for the increased risk of admission for critical care in the VOC B.1.1.7 versus the non VOC B.1.1.7 group (adjusted HR 1.28 95% CI: 1.05 - 1.56). A sensitivity analysis, including only those patients with a minimum of 20-days follow-up was consistent with the main analysis (adjusted HR: 2.08; 95% CI: 1.56-2.78) (the characteristics of the primary care cohort restricted to patients with a minimum of 20-days of follow-up are presented in supplementary Table S2).

**Figure 3:**
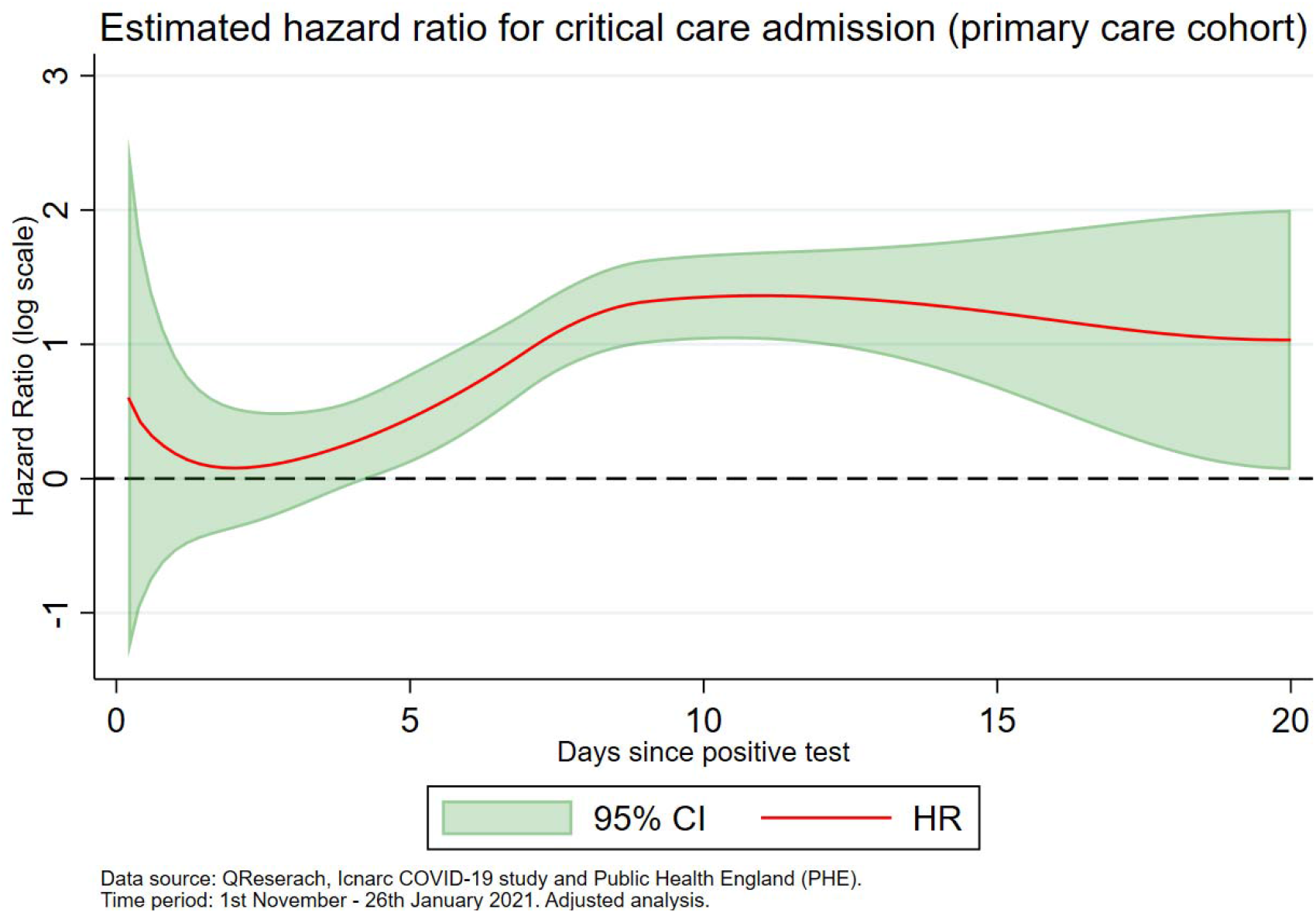
Estimated adjusted hazard ratio for critical care admission for primary care patients tested positive in the community between 1^st^ November 2020 and 26^th^ January 2021 (primary care cohort). The dotted black line is equal to 1.

The characteristics of patients who died with COVID-19 and of those who were admitted for critical care compared with those alive and not admitted, respectively, are summarised in supplementary Table S3.

### Critical care cohort

During the study period (1 November 2020 to 27 January 2021), there were 2,115,220 positive COVID-19 RT-PCR tests from PHE and 13,919 admissions for critical care in the ICNARC COVID-19 study. Combined, this produced a linked dataset of 13,402 patients, of which, 6,040 had a positive COVID-19 RT-PCR test in the community (not hospital). SGTF status (as a proxy for VOC B.1.1.7 B.1.1.7) was identifiable in 3,432 (56.8 %) (Figure 1).

#### VOC B.1.1.7

Of the 3,432 patients for whom results were available, 2,019 (58.8%) had VOC B.1.1.7 and 1,413 (42.2%) had non-VOC B.1.1.7 (Figure 1). Figure 2 shows the Kaplan-Meier plot for risk of admission for critical care, by variant, for the complete case analysis.

Table 2 compares the demographic characteristics of the patients in the critical care cohort in the VOC B.1.1.7 group compared with the non-VOC B.1.1.7 group. Patients in the VOC B.1.1.7 group tended to be marginally younger (means 57.8 versus 59.3 years) and less likely to have a higher BMI than those in the non-VOC B.1.1.7 group. Acute severity of illness, as measured by the APACHE II score, tended to be lower in the VOC B.1.1.7 group, but the proportion receiving invasive mechanical ventilation, within the first 24 hours of critical care, was similar. There were 1173/2019 (58.1%) of the VOC B.1.1.7 group who had completed their critical care stay at the point of analysis, compared with 1235/1413 (87.4%) of the non-VOC B.1.1.7 group (Supplementary Table S6).

**Table 2:**
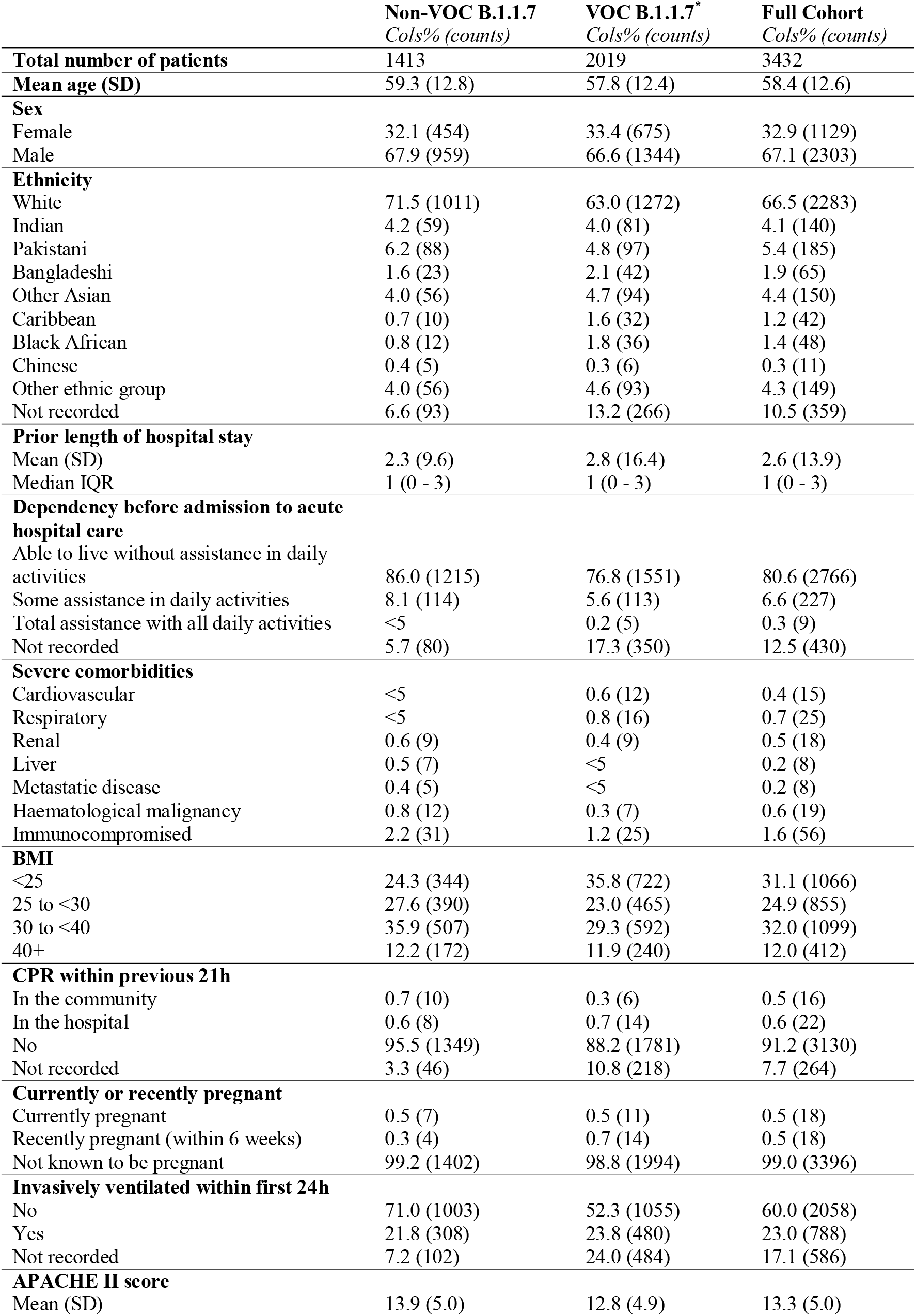
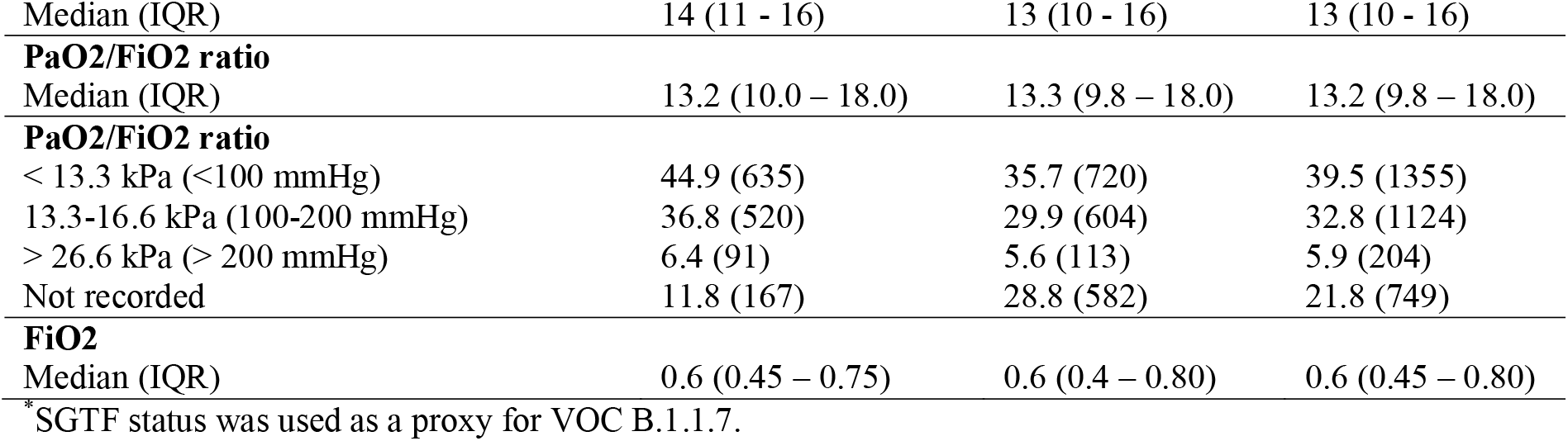
Demographic, medical characteristics and indicators of acute severity observed for critically ill patients tested positive in the community between 1^st^ November 2020 and 27^th^ January 2021 (critical care cohort), by variant.

#### Risk of mortality at the end of critical care

A lower risk of admission for critical care in the VOC B.1.1.7 group, in the unadjusted analysis (unadjusted HR: 0.79, 95% CI: 0.67 - 0.93), was mainly accounted for after adjustment for date of admission to critical care (HR: 0.84, 95% CI: 0.64 - 0.99). After adjusting for additional confounders, critical care mortality did not differ significantly between the VOC B.1.1.7 and non-VOC B.1.1.7 group (adjusted HR: 0.93, 95% CI 0.76-1.15). We found no evidence of a significant interaction between VOC B.1.1.7 and: ethnic group (p = 0.68); age group (p = 0.60) or sex (p = 0.86). Sensitivity analyses, including only those already discharged from critical care (alive or dead), were consistent with the main analysis (characteristics of the critical care cohort restricted to those who had completed their critical care stay are summarised in supplementary Table S4).

Complete case analyses were all consistent with the imputed analyses above.

#### Types and duration of organ support

Table 3 shows the outcomes and organ support for patients in the matched cohort. Overall, organ support receipt was similar between the two groups. Demographic and clinical characteristics of patients in the matched cohort are shown in Table S4 and, of those with complete outcomes (death or survival at discharge from critical care) are summarised in Tables S5-S6.

**Tables 3:**
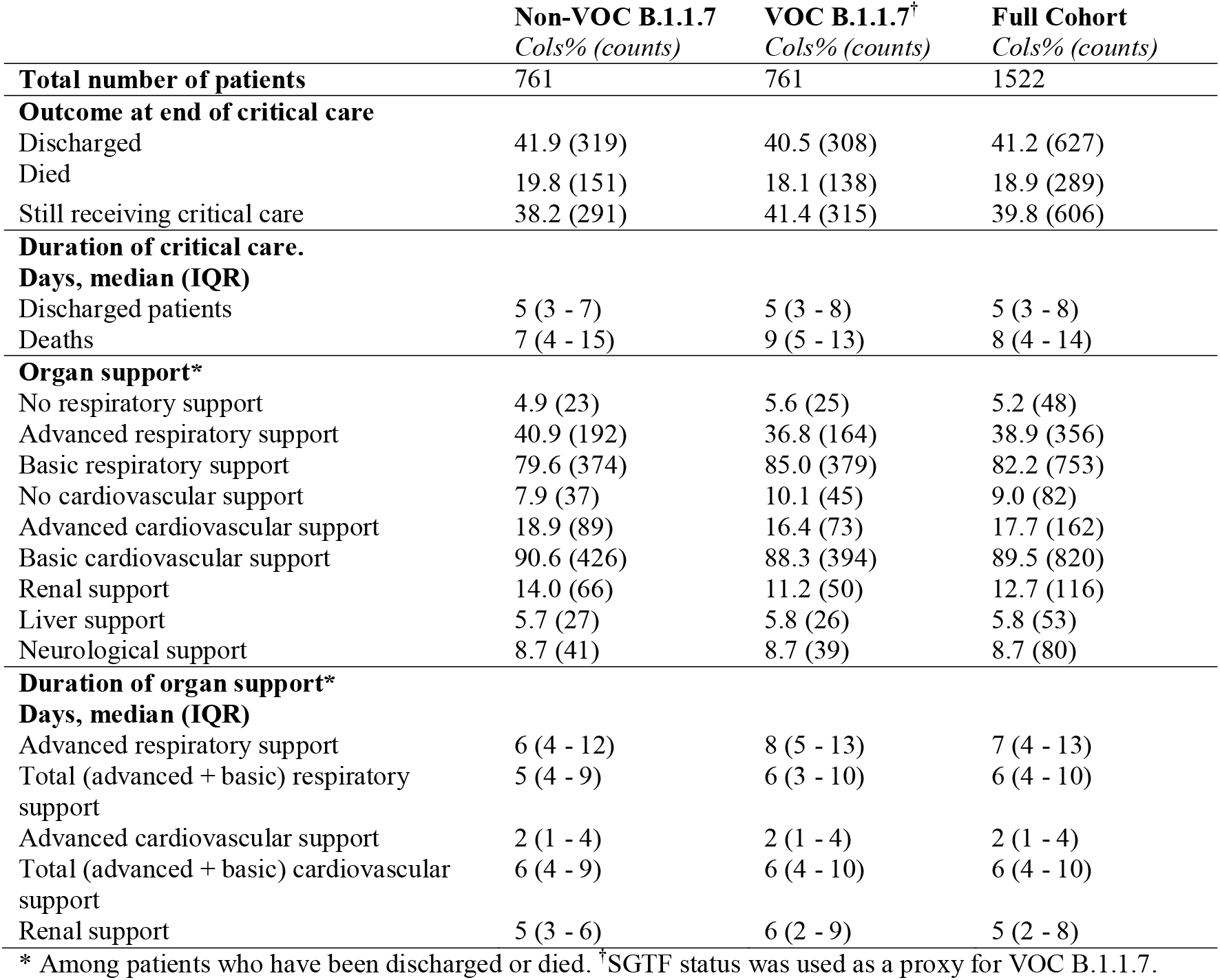
Critical care outcomes for the matched cohort of critically ill patients tested positive in the community between 1^st^ November 2020 and 27th January 2021, by variant. Each patient with VOC B.1.1.7 was matched with a patient without VOC B.1.1.7 admitted to the same ICU unit. Only pair of patients who were admitted within 3 days of each other. For each patient with VOC B.1.1.7 only one matched set was randomly selected. The matched cohort consists of 1522 patients (761 in the VOC B.1.1.7 group and 761 in the non-VOC B.1.1.7 group).

|  | **Non-VOC B.1.1.7**<br>*Cols% (counts)* | **VOC B.1.1.7<sup>†</sup>**<br>*Cols% (counts)* | **Full Cohort**<br>*Cols% (counts)* |
| --- | --- | --- | --- |
| **Total number of patients** | 761 | 761 | 1522 |
| **Outcome at end of critical care** |  |  |  |
| Discharged | 41.9 (319) | 40.5 (308) | 41.2 (627) |
| Died | 19.8 (151) | 18.1 (138) | 18.9 (289) |
| Still receiving critical care | 38.2 (291) | 41.4 (315) | 39.8 (606) |
| **Duration of critical care.** |  |  |  |
| **Days, median (IQR)** |  |  |  |
| Discharged patients | 5 (3 - 7) | 5 (3 - 8) | 5 (3 - 8) |
| Deaths | 7 (4 - 15) | 9 (5 - 13) | 8 (4 - 14) |
| **Organ support\*** |  |  |  |
| No respiratory support | 4.9 (23) | 5.6 (25) | 5.2 (48) |
| Advanced respiratory support | 40.9 (192) | 36.8 (164) | 38.9 (356) |
| Basic respiratory support | 79.6 (374) | 85.0 (379) | 82.2 (753) |
| No cardiovascular support | 7.9 (37) | 10.1 (45) | 9.0 (82) |
| Advanced cardiovascular support | 18.9 (89) | 16.4 (73) | 17.7 (162) |
| Basic cardiovascular support | 90.6 (426) | 88.3 (394) | 89.5 (820) |
| Renal support | 14.0 (66) | 11.2 (50) | 12.7 (116) |
| Liver support | 5.7 (27) | 5.8 (26) | 5.8 (53) |
| Neurological support | 8.7 (41) | 8.7 (39) | 8.7 (80) |
| **Duration of organ support\*** |  |  |  |
| **Days, median (IQR)** |  |  |  |
| Advanced respiratory support | 6 (4 - 12) | 8 (5 - 13) | 7 (4 - 13) |
| Total (advanced + basic) respiratory support | 5 (4 - 9) | 6 (3 - 10) | 6 (4 - 10) |
| Advanced cardiovascular support | 2 (1 - 4) | 2 (1 - 4) | 2 (1 - 4) |
| Total (advanced + basic) cardiovascular support | 6 (4 - 9) | 6 (4 - 10) | 6 (4 - 10) |
| Renal support | 5 (3 - 6) | 6 (2 - 9) | 5 (2 - 8) |

\* Among patients who have been discharged or died. <sup>†</sup>SGTF status was used as a proxy for VOC B.1.1.7.

## DISCUSSION

To our knowledge this is the first study to report risks of admission to critical care and clinical outcomes among patients admitted to critical care comparing VOC B.1.1.7 with the non-VOC B1.1.7 variant. VOC B.1.1.7 became dominant over the study period. Using SGTF results, as a proxy for VOC B.1.1.7, we found a substantially increased risk of overall COVID-19 28-day mortality and of admission for critical care associated with VOC B.1.1.7 but no difference in risk of critical care mortality or organ support receipt for patients in critical care.

We found a 60% higher risk of COVID-19 28-day mortality for people with VOC B.1.1.7 compared with those in the non-VOC B.1.1.7 group. Our study shows that the highly prevalent VOC B.1.1.7 infects a similar population to the non-VOC B1.1.7 variant, albeit with fewer patients aged 70 years and older having a positive community test for VOC B.1.1.7. Whether this is a true difference in who the variant infects, or a difference in exposure or in testing remains unclear. An advantage of our study is that the primary care cohort had prior recording of a wide range of exposures and co-morbidities. This allowed us to adjust the analysis, controlling for many important, potential confounders. This adjustment determined the increased risk of COVID-19 28-day mortality (mainly explained by adjustment for age) which was not obvious in the unadjusted analysis.

Infection with VOC B.1.1.7 (VOC B.1.1.7 group) was associated with a doubling of the risk of admission for critical care compared with the non-VOC B.1.1.7 group. Although the increased infectivity of VOC B.1.1.7 has been reported ^5-6^, we are not aware of prior work examining the risk of critical care admission. Adjusting only for the date of positive test, did not explain this increased risk of admission for critical care, suggesting that the effect is not explained by time-dependent factors such as critical care bed availability. Although the finding raises concerns about future capacity planning, it should be interpreted with some caution. As an example, in our study population VOC B.1.1.7 appears to be more prevalent in younger people and we do not know, with the data available, whether this will remain the case as VOC B.1.1.7 spreads. The large preponderance of men admitted for critical care, despite fewer men having a positive COVID-19 RT-PCR test, has been previously reported^17^. Our work shows this pattern remains with VOC B.1.1.7. Both findings are similar to those reported for UK data for all critical care admissions, where males predominate ^20^. The finding that, once admitted for critical care, outcomes are similar between those in the VOC B.1.1.7 and non-VOC B.1.1.7 group, may result from patients in either group having similar acute severity of illness at the point of admission.

Our study has some important strengths. It uses established, complete and validated data sources which are either the national databases for England (PHE and ICNARC COVID-19 study) or a very large representative sample (QResearch) ^17,18,21,22^. Therefore, our results are likely to be generalisable. Outcome data, for both patient cohorts, were complete and missing data occurred only in some predictor variables. Multiple imputation agreed with complete case analyses in both cohorts. We restricted our analysis to patients with community, laboratory-confirmed, positive test results in order to be able to identify VOC B.1.1.7 using the SGTF proxy, which has proven to be a good proxy for monitoring trends in VOC B.1.1.7. Use of the national register also minimises the risk of misclassification bias, however, misclassification bias may still have occurred. A significant limitation of the data available for our work is that determination of SGTF status, as a proxy for VOC B.1.1.7, was only possible in just over 50% of patients with a community COVID-19 RT-PCR positive test. To minimise the risks of changing eligibility over time, we restricted our analysis to an eleven-week period where VOC B.1.1.7 had become significant. We used established measures, including critical care mortality and measures of critical care severity, including duration of critical care stay and receipt of basic and advanced organ support. Our data are very timely, being reported during the third pandemic wave in England. However, due to the data available, we were able to estimate risk of critical care admission and mortality only among individuals tested in the community, who are generally healthier and younger than those tested in hospitals. More robust results will be possible as more testing data becomes available. As with all observational studies, our study remains subject to unmeasured confounding.

Our study demonstrates increased risk of COVID-19 28-day mortality and risk of critical care admission for patients who test positive for VOC B.1.1.7. Combined with evidence of increased infectivity, our findings emphasise the importance of measures to control exposure to and infection with COVD-19.

## Data Availability

To guarantee the confidentiality of personal and health information only the authors have had access to the data during the study in accordance with the relevant licence agreements. Access to the QResearch data is according to the information on the QResearch website (www.qresearch.org). The equations presented in this paper will be released as Open Source Software under the GNU lesser GPL v3 to ensure transparency and wide availability. The open source software allows use without charge.

## Acknowledgements

This project involves data derived from patient-level information collected by the NHS, as part of the care and support of patients. We acknowledge the contribution of EMIS (Egton Medical Information Systems) practices who contribute to the QResearch database and EMIS, and the Universities of Nottingham and Oxford for expertise in establishing, developing, and supporting the QResearch database. QResearch acknowledges funding from the NIHR funded Nottingham Biomedical Research Centre. Access to the data was facilitated by the PHE Office for Data Release, ICNARC and the QResearch Scientific Committee at the University of Oxford.

## Authors and contributors

Study conceptualisation was led by MP, JHC, PW. All authors contributed to the development of the research question, study design, with development of advanced statistical aspects led by MP, CC, KT and JHC. MP led the statistical analyses which were checked by WL. All authors contributed to the interpretation of the results. JHC, PW and MP wrote the first draft of the paper. All authors contributed to the critical revision of the manuscript for important intellectual content and approved the final version of the manuscript.

MP, JHC and WL had full access to all data in the study and takes responsibility of the integrity of the rate, and transparent account of the study being reported; that no important aspects of the study have been omitted; and that any discrepancies from the study as planned have been explained.

## Ethics approval

The ethics approval for the development and validation of QCovid was granted by the East Midlands-Derby Research Ethics Committee [reference 18/EM/0400].

## Funding

This study was supported by funds from the Wellcome Trust (grant number is 221514/Z/20/Z), NIHR Oxford Biomedical Research Centre and the Medical Sciences Division of the University of Oxford. QResearch is funded by the John Fell Oxford University Press Research Fund, the Oxford Wellcome Institutional Strategic Support Fund (204826/Z/16/Z), Cancer Research UK (CR-UK) grant number C5255/A18085, through the Cancer Research UK Oxford Centre and It also receives contributions in kind from EMIS Health (commercial supplier of NHS clinical computer systems).

## Role of the funding source

The funders external to the University of Oxford, had no part in study design; in the collection, analysis, and interpretation of data; in the writing of the report; nor in the decision to submit the paper for publication.

## Competing interests

JHC reports grants from the Wellcome trust, HDR-UK, National Institute for Health Research Biomedical Research Centre, Oxford, grants from John Fell Oxford University Press Research Fund, grants from Cancer Research UK (CR-UK) grant number C5255/A18085, through the Cancer Research UK Oxford Centre, grants from the Oxford Wellcome Institutional Strategic Support Fund (204826/Z/16/Z), JHC is a member of SAGE subgroups on ethnicity and chair of the NERVTAG risk stratification subgroup. JHC is an unpaid director of QResearch, a not-for-profit organisation which is a partnership between the University of Oxford and EMIS Health who supply the QResearch database used for this work. JHC is founder and former director of ClinRisk Ltd outside the submitted work. PST reports previous consultations with AstraZeneca and Duke-NUS outside the submitted work. PW was Chief Medical Officer for Sensyne Health and his department received research funding from Sensyne Health. He holds shares in the company. He received grant funding from the National Institute for Health Research and Wellcome.

## Declaration

The lead author affirms that the manuscript is an honest, accurate, and transparent account of the study being reported; that no important aspects of the study have been omitted and that any discrepancies from the study as planned have been explained.

