## Supplemental material for "Analysis of severe outcomes associated with the SARS-CoV-2 Variant of Concern 202012/01 in England using ICNARC Case Mix Programme and QResearch databases"

Martina Patone PhD ^1^

Karen Thomas^2^

Rob Hatch^3^

Pui San Tan PhD^4^

Carol Coupland PhD^5^

Weiqi Liao^6^

Paul Mouncey^7^

David Harrison PhD^8^

Kathryn Rowan PhD^9^

Peter Horby FRCP^10^

¥ Peter Watkinson^11^

¥Julia Hippisley-Cox FRCP^12^

**Institutions**

^1^Statistician, Nuffield Department of Primary Care Health Sciences, University of Oxford

^2^Senior Statistician, Intensive Care National Audit & Research Centre

3 Clinical Research Fellow, Nuffield Department of Clinical Neurosciences, University of Oxford

^4^Data Scientist and Pharmacist, Nuffield Department of Primary Care Health Sciences, University of Oxford

^5^Professor of Medical Statistics in Primary Care, Division of Primary Care, School of Medicine, University of Nottingham

^6^ Data Scientist, Nuffield Department of Primary Care Health Sciences, University of Oxford

^7^Head of Research, Intensive Care National Audit & Research Centre

^8^Head Statistician, Intensive Care National Audit & Research Centre and Honorary Professor, Department of Medical Statistics, London School of Hygiene and Tropical Medicine

^9^Founder/Director, Intensive Care National Audit & Research Centre and Honorary Professor, Department of Health Services Research and Policy, London School of Hygiene & Tropical Medicine

^10^Professor of Emerging Infectious Diseases, University of Oxford

^11^ Associate Professor of Intensive Care Medicine, Consultant in Intensive Care Medicine, NIHR Biomedical Research Centre, Oxford, Nuffield Department of Clinical Neurosciences, University of Oxford, Oxford University Hospitals NHS Trust

^12^Professor of Clinical Epidemiology and General Practice, Nuffield Department of Primary Care Health Sciences, University of Oxford

¥ joint senior authors

**Author for correspondence**

**Supplementary material**

**Table S1: Demographics of primary care patients tested positive in the community between 1^st^ November 2020 and 26^th^ January 2021 (primary care cohort) who have a minimum of 28 days of follow up from the day of their positive test, by variant.**

|  | **Non-VOC B.1.1.7** | **VOC B.1.1.7^*^** | **Full Cohort** |
| --- | --- | --- | --- |
|  | *Cols% (counts)* | *Cols% (counts)* | *Cols% (counts)* |
| **Total number of patients** | 66756 | 43583 | 110339 |
| **ICU admitted** | 0.3 (225) | 0.4 (184) | 0.4 (409) |
| **Deaths** | 0.3 (185) | 0.1 (58) | 0.2 (243) |
| **Sex** |  |  |  |
| Female | 53.3 (35579) | 52.7 (22956) | 53.1 (58535) |
| Male | 46.7 (31177) | 47.3 (20627) | 46.9 (51804) |
| **Mean age (SD)** | 37.6 (18.1) | 35.9 (17.4) | 36.9 (17.8) |
| **Age Categories** |  |  |  |
| 18-29 | 36.9 (24633) | 38.1 (16612) | 37.4 (41245) |
| 30-39 | 19.1 (12781) | 20.2 (8806) | 19.6 (21587) |
| 40-49 | 16.6 (11067) | 18.4 (7999) | 17.3 (19066) |
| 50-59 | 15.3 (10227) | 14.3 (6242) | 14.9 (16469) |
| 60-69 | 7.6 (5051) | 6.2 (2699) | 7.0 (7750) |
| 70-79 | 3.0 (2010) | 2.1 (903) | 2.6 (2913) |
| 80-89 | 1.2 (799) | 0.6 (269) | 1.0 (1068) |
| 90-99 | 0.3 (188) | 0.1 (53) | 0.2 (241) |
| **Ethnicity** |  |  |  |
| White | 60.4 (40309) | 59.4 (25906) | 60.0 (66215) |
| Indian | 3.9 (2620) | 4.3 (1888) | 4.1 (4508) |
| Pakistani | 5.4 (3573) | 3.2 (1400) | 4.5 (4973) |
| Bangladeshi | 2.7 (1813) | 3.5 (1527) | 3.0 (3340) |
| Other Asian | 2.0 (1340) | 2.6 (1123) | 2.2 (2463) |
| Caribbean | 0.5 (327) | 1.0 (446) | 0.7 (773) |
| Black African | 1.7 (1105) | 2.2 (957) | 1.9 (2062) |
| Chinese | 0.2 (137) | 0.3 (146) | 0.3 (283) |
| Other ethnic group | 3.4 (2243) | 4.7 (2060) | 3.9 (4303) |
| Not recorded | 19.9 (13289) | 18.7 (8130) | 19.4 (21419) |
| **Date of positive test** |  |  |  |
| 1 Nov to 14 Nov | 39.2 (26160) | 4.1 (1784) | 25.3 (27944) |
| 15 Nov to 28 Nov | 24.7 (16509) | 6.6 (2879) | 17.6 (19388) |
| 29 Nov to 12 Dec | 16.5 (10990) | 17.7 (7704) | 16.9 (18694) |
| 13 Dec to 26 Dec | 16.2 (10847) | 54.4 (23700) | 31.3 (34547) |
| 27 Dec to 10 Jan | 3.4 (2250) | 17.2 (7516) | 8.9 (9766) |
| **House size** |  |  |  |
| 1 person | 26.4 (17639) | 23.7 (10324) | 25.3 (27963) |
| 2 people | 20.3 (13559) | 18.4 (8010) | 19.5 (21569) |
| 3-5 people | 45.3 (30253) | 50.3 (21904) | 47.3 (52157) |
| 6+ people | 7.9 (5305) | 7.7 (3345) | 7.8 (8650) |
| **House type** |  |  |  |
| Neither | 99.7 (66548) | 99.8 (43489) | 99.7 (110037) |
| Carehome | 0.3 (170) | 0.1 (61) | 0.2 (231) |
| Homeless | 0.1 (38) | 0.1 (33) | 0.1 (71) |
| **BMI** |  |  |  |
| <25 | 54.2 (36205) | 54.9 (23940) | 54.5 (60145) |
| 25-30 | 13.1 (8713) | 11.3 (4928) | 12.4 (13641) |
| 30-40 | 5.5 (3667) | 4.6 (1986) | 5.1 (5653) |
| >= 40 | 3.1 (2086) | 2.3 (984) | 2.8 (3070) |
| Not recorded | 24.1 (16085) | 26.9 (11745) | 25.2 (27830) |
| **Smoking status** |  |  |  |
| Non smoker | 57.5 (38381) | 55.4 (24164) | 56.7 (62545) |
| Ex smoker | 17.3 (11580) | 16.8 (7306) | 17.1 (18886) |
| Light smoker | 7.7 (5150) | 8.1 (3529) | 7.9 (8679) |
| Moderate smoker | 1.4 (953) | 1.3 (550) | 1.4 (1503) |
| Heavy smoker | 0.5 (317) | 0.4 (191) | 0.5 (508) |
| Not recorded | 15.5 (10375) | 18.0 (7843) | 16.5 (18218) |
| **Geographical region** |  |  |  |
| East Midlands | 2.0 (1317) | 0.8 (342) | 1.5 (1659) |
| East of England | 1.8 (1215) | 3.5 (1509) | 2.5 (2724) |
| London | 18.1 (12115) | 43.0 (18721) | 27.9 (30836) |
| North East | 5.0 (3364) | 0.9 (403) | 3.4 (3767) |
| North West | 30.7 (20480) | 7.0 (3033) | 21.3 (23513) |
| South Central | 8.7 (5814) | 12.1 (5276) | 10.1 (11090) |
| South East | 4.9 (3250) | 24.1 (10509) | 12.5 (13759) |
| South West | 5.0 (3311) | 2.8 (1201) | 4.1 (4512) |
| West Midlands | 17.8 (11853) | 5.3 (2300) | 12.8 (14153) |
| Yorkshire & Humber | 6.0 (4037) | 0.7 (289) | 3.9 (4326) |
| **Deprivation quintile** |  |  |  |
| 1 (least deprived) | 21.9 (14646) | 19.1 (8304) | 20.8 (22950) |
| 2 | 21.6 (14428) | 22.3 (9701) | 21.9 (24129) |
| 3 | 21.1 (14111) | 21.1 (9164) | 21.1 (23275) |
| 4 | 19.9 (13301) | 19.0 (8286) | 19.6 (21587) |
| 5 (most deprived) | 14.7 (9793) | 18.0 (7856) | 16.0 (17649) |
| Not recorded | 0.7 (477) | 0.7 (272) | 0.7 (749) |
| **Comorbidities** |  |  |  |
| Asthma | 15.5 (10375) | 14.4 (6268) | 15.1 (16643) |
| COPD | 1.0 (683) | 0.6 (277) | 0.9 (960) |
| Diabetes type 1 | 0.6 (378) | 0.5 (224) | 0.5 (602) |
| Diabetes type 2 | 5.0 (3344) | 3.7 (1599) | 4.5 (4943) |
| Hypertension | 10.3 (6847) | 8.1 (3550) | 9.4 (10397) |
| Parkinson | 0.1 (50) | 0.1 (26) | 0.1 (76) |
| Epilepsy | 1.1 (713) | 0.9 (407) | 1.0 (1120) |
| Cerebral palsy | 0.1 (60) | 0.1 (33) | 0.1 (93) |
| Motor Neuron Disease | <5 | <5 | 0.0 (6) |
| Huntington’s disease | <5 | <5 | <5 |
| Myasthenia | 0.1 (92) | 0.1 (50) | 0.1 (142) |
| Downs syndrome | 0.0 (19) | 0.0 (8) | 0.0 (27) |
| Learning disabilities excluding downs | 0.0 (22) | 0.0 (14) | 0.0 (36) |

^*^SGTF status was used as a proxy for VOC B.1.1.7.

**Table S2: Demographic characteristics observed for primary care patients tested positive in the community between 1^st^ November 2020 and 26^th^ January 2021 (primary-care cohort) who have a minimum of 20 days of follow up from the day of their positive test, by variant.**

|  | **Non-VOC B.1.1.7** | **VOC B.1.1.7^*^** | **Full Cohort** |
| --- | --- | --- | --- |
|  | *Cols% (counts)* | *Cols% (counts)* | *Cols% (counts)* |
| **Total number of patients** | 73085 | 69580 | 142665 |
| **ICU admitted** | 0.3 (243) | 0.4 (299) | 0.4 (542) |
| **Deaths** | 0.4 (270) | 0.4 (248) | 0.4 (518) |
| **Sex** |  |  |  |
| Female | 53.3 (38951) | 52.5 (36512) | 52.9 (75463) |
| Male | 46.7 (34134) | 47.5 (33068) | 47.1 (67202) |
| **Mean age (SD)** | 37.8 (18.1) | 36.7 (17.3) | 37.2 (17.7) |
| **Age Categories** |  |  |  |
| 18-29 | 36.6 (26717) | 36.9 (25708) | 36.7 (52425) |
| 30-39 | 19.2 (14037) | 20.3 (14136) | 19.7 (28173) |
| 40-49 | 16.5 (12086) | 18.0 (12493) | 17.2 (24579) |
| 50-59 | 15.5 (11299) | 14.8 (10285) | 15.1 (21584) |
| 60-69 | 7.7 (5627) | 6.8 (4750) | 7.3 (10377) |
| 70-79 | 3.0 (2229) | 2.4 (1647) | 2.7 (3876) |
| 80-89 | 1.2 (885) | 0.7 (482) | 1.0 (1367) |
| 90-99 | 0.3 (205) | 0.1 (79) | 0.2 (284) |
| **Ethnicity** |  |  |  |
| White | 60.6 (44323) | 58.8 (40943) | 59.8 (85266) |
| Indian | 3.9 (2837) | 4.3 (2973) | 4.1 (5810) |
| Pakistani | 5.2 (3794) | 3.2 (2259) | 4.2 (6053) |
| Bangladeshi | 2.6 (1909) | 3.4 (2379) | 3.0 (4288) |
| Other Asian | 2.0 (1426) | 2.5 (1758) | 2.2 (3184) |
| Caribbean | 0.5 (365) | 1.1 (787) | 0.8 (1152) |
| Black African | 1.6 (1188) | 2.3 (1579) | 1.9 (2767) |
| Chinese | 0.2 (145) | 0.3 (234) | 0.3 (379) |
| Other ethnic group | 3.3 (2439) | 4.7 (3281) | 4.0 (5720) |
| Not recorded | 20.1 (14659) | 19.2 (13387) | 19.7 (28046) |
| **Date of positive test** |  |  |  |
| 1 Nov to 14 Nov | 35.8 (26160) | 2.6 (1784) | 19.6 (27944) |
| 15 Nov to 28 Nov | 22.6 (16509) | 4.1 (2879) | 13.6 (19388) |
| 29 Nov to 12 Dec | 15.0 (10990) | 11.1 (7704) | 13.1 (18694) |
| 13 Dec to 26 Dec | 14.8 (10847) | 34.1 (23700) | 24.2 (34547) |
| 27 Dec to 10 Jan | 11.7 (8579) | 48.2 (33513) | 29.5 (42092) |
| **House size** |  |  |  |
| 1 person | 26.6 (19452) | 24.6 (17136) | 25.6 (36588) |
| 2 people | 20.4 (14938) | 19.0 (13214) | 19.7 (28152) |
| 3-5 people | 45.2 (33053) | 48.7 (33851) | 46.9 (66904) |
| 6+ people | 7.7 (5642) | 7.7 (5379) | 7.7 (11021) |
| **House type** |  |  |  |
| Neither | 99.7 (72866) | 99.8 (69441) | 99.7 (142307) |
| Carehome | 0.2 (177) | 0.1 (90) | 0.2 (267) |
| Homeless | 0.1 (42) | 0.1 (49) | 0.1 (91) |
| **BMI** |  |  |  |
| <25 | 54.4 (39770) | 55.5 (38649) | 55.0 (78419) |
| 25-30 | 13.1 (9608) | 11.8 (8221) | 12.5 (17829) |
| 30-40 | 5.5 (4025) | 4.7 (3298) | 5.1 (7323) |
| >= 40 | 3.1 (2270) | 2.4 (1657) | 2.8 (3927) |
| Not recorded | 23.8 (17412) | 25.5 (17755) | 24.7 (35167) |
| **Smoking status** |  |  |  |
| Non smoker | 57.6 (42077) | 55.9 (38901) | 56.8 (80978) |
| Ex smoker | 17.5 (12821) | 17.2 (11943) | 17.4 (24764) |
| Light smoker | 7.8 (5688) | 8.8 (6107) | 8.3 (11795) |
| Moderate smoker | 1.5 (1060) | 1.4 (993) | 1.4 (2053) |
| Heavy smoker | 0.5 (360) | 0.5 (342) | 0.5 (702) |
| Not recorded | 15.2 (11079) | 16.2 (11294) | 15.7 (22373) |
| **Geographical region** |  |  |  |
| East Midlands | 1.9 (1410) | 0.9 (632) | 1.4 (2042) |
| East of England | 1.8 (1338) | 3.5 (2438) | 2.6 (3776) |
| London | 17.5 (12787) | 38.7 (26927) | 27.8 (39714) |
| North East | 5.1 (3748) | 1.4 (979) | 3.3 (4727) |
| North West | 31.5 (23046) | 10.9 (7592) | 21.5 (30638) |
| South Central | 8.5 (6223) | 12.2 (8517) | 10.3 (14740) |
| South East | 4.7 (3442) | 19.9 (13875) | 12.1 (17317) |
| South West | 5.0 (3657) | 3.2 (2194) | 4.1 (5851) |
| West Midlands | 17.9 (13060) | 8.2 (5734) | 13.2 (18794) |
| Yorkshire & Humber | 6.0 (4374) | 1.0 (692) | 3.6 (5066) |
| **Deprivation quintile** |  |  |  |
| 1 (least deprived) | 21.9 (16034) | 19.5 (13548) | 20.7 (29582) |
| 2 | 21.7 (15888) | 22.1 (15386) | 21.9 (31274) |
| 3 | 21.2 (15529) | 20.9 (14538) | 21.1 (30067) |
| 4 | 19.8 (14508) | 18.9 (13214) | 19.4 (27722) |
| 5 (most deprived) | 14.5 (10607) | 17.8 (12418) | 16.2 (23025) |
| Not recorded | 0.7 (519) | 0.7 (476) | 0.7 (995) |
| **Comorbidities** |  |  |  |
| Asthma | 15.6 (11395) | 14.5 (10090) | 15.1 (21485) |
| COPD | 1.0 (762) | 0.7 (477) | 0.9 (1239) |
| Diabetes type 1 | 0.6 (409) | 0.5 (359) | 0.5 (768) |
| Diabetes type 2 | 5.0 (3657) | 3.9 (2718) | 4.5 (6375) |
| Hypertension | 10.3 (7560) | 8.6 (5997) | 9.5 (13557) |
| Parkinson | 0.1 (56) | 0.1 (39) | 0.1 (95) |
| Epilepsy | 1.1 (782) | 0.9 (655) | 1.0 (1437) |
| Cerebral palsy | 0.1 (64) | 0.1 (50) | 0.1 (114) |
| Motor Neuron Disease | <5 | <5 | 0.0 (7) |
| Huntington’s disease | <5 | <5 | <5 |
| Myasthenia | 0.1 (99) | 0.1 (88) | 0.1 (187) |
| Downs syndrome | 0.0 (20) | 0.0 (14) | 0.0 (34) |
| Learning disabilities excluding downs | 0.0 (25) | 0.0 (28) | 0.0 (53) |

^*^SGTF status was used as a proxy for VOC B.1.1.7.

**Table S3: Demographic and clinical characteristics for primary care patients tested positive in the community between 1^st^ November 2020 and 26^th^ January 2021 (primary care cohort) who survived or died within the study period and those who have received or not critical care.**

|  | **Alive** | **Died** | **Not critical care** | **Critical care** |
| --- | --- | --- | --- | --- |
|  | *Cols% (counts)* | *Cols% (counts)* | *Cols% (counts)* | *Cols% (counts)* |
| **Total number of patients** | 197807 | 613 | 197708 | 712 |
| **ICU admitted/Deaths** | 0.3 (563) | 24.3 (149) | 0.2 (493) | 24.9 (177) |
| **Sex** |  |  |  |  |
| Female | 52.8 (104415) | 39.2 (240) | 52.8 (104428) | 31.9 (227) |
| Male | 47.2 (93392) | 60.8 (373) | 47.2 (93280) | 68.1 (485) |
| **Mean age (SD)** | 37.5 (17.7) | 74.7 (14.6) | 37.6 (17.8) | 57.9 (12.4) |
| **Age Categories** |  |  |  |  |
| 18-29 | 36.0 (71121) | <5 | 36.0 (71110) | 1.7 (12) |
| 30-39 | 19.9 (39411) | 2.1 (13) | 19.9 (39377) | 6.6 (47) |
| 40-49 | 17.1 (33858) | 3.1 (19) | 17.1 (33769) | 15.2 (108) |
| 50-59 | 15.6 (30879) | 10.9 (67) | 15.5 (30740) | 28.9 (206) |
| 60-69 | 7.5 (14875) | 17.1 (105) | 7.5 (14771) | 29.4 (209) |
| 70-79 | 2.7 (5359) | 23.7 (145) | 2.7 (5392) | 15.7 (112) |
| 80-89 | 1.0 (1933) | 26.4 (162) | 1.1 (2077) | 2.5 (18) |
| 90-99 | 0.2 (371) | 16.5 (101) | 0.2 (472) | 0 |
| **Ethnicity** |  |  |  |  |
| White | 59.4 (117530) | 64.1 (393) | 59.4 (117485) | 61.5 (438) |
| Indian | 4.0 (7918) | 4.6 (28) | 4.0 (7900) | 6.5 (46) |
| Pakistani | 4.2 (8336) | 5.2 (32) | 4.2 (8331) | 5.2 (37) |
| Bangladeshi | 2.8 (5547) | 1.6 (10) | 2.8 (5543) | 2.0 (14) |
| Other Asian | 2.3 (4476) | 1.3 (8) | 2.3 (4469) | 2.1 (15) |
| Caribbean | 0.9 (1729) | 1.6 (10) | 0.9 (1727) | 1.7 (12) |
| Black African | 2.1 (4091) | 1.1 (7) | 2.1 (4080) | 2.5 (18) |
| Chinese | 0.3 (529) | 0.5 (3) | 0.3 (530) | <5 |
| Other ethnic group | 3.9 (7785) | 3.9 (24) | 3.9 (7776) | 4.6 (33) |
| Not recorded | 20.2 (39866) | 12.6 (77) | 20.2 (39867) | 13.6 (97) |
| **Date of positive test** |  |  |  |  |
| 1 Nov to 14 Nov | 14.1 (27853) | 14.8 (91) | 14.1 (27848) | 13.5 (96) |
| 15 Nov to 28 Nov | 9.8 (19335) | 8.6 (53) | 9.8 (19317) | 10.0 (71) |
| 29 Nov to 12 Dec | 9.4 (18637) | 9.3 (57) | 9.4 (18627) | 9.4 (67) |
| 13 Dec to 26 Dec | 17.4 (34409) | 22.5 (138) | 17.4 (34420) | 17.8 (127) |
| 27 Dec to 10 Jan | 29.4 (58092) | 32.1 (197) | 29.4 (58029) | 36.5 (260) |
| 11 Jan to 26 Jan | 20.0 (39481) | 12.4 (76) | 20.0 (39467) | 12.8 (91) |
| **House size** |  |  |  |  |
| 1 person | 26.2 (51781) | 37.7 (231) | 26.2 (51784) | 32.0 (228) |
| 2 people | 20.1 (39798) | 29.0 (178) | 20.1 (39802) | 24.4 (174) |
| 3-5 people | 46.1 (91260) | 19.2 (118) | 46.1 (91117) | 36.7 (261) |
| 6+ people | 7.6 (14968) | 14.0 (86) | 7.6 (15005) | 6.9 (49) |
| **House type** |  |  |  |  |
| Neither | 99.7 (197205) | 91.8 (563) | 99.7 (197057) | 99.9 (711) |
| Carehome | 0.2 (447) | 8.2 (50) | 0.3 (496) | <5 |
| Homeless | 0.1 (155) | 0 | 0.1 (155) | 0 |
| **BMI** |  |  |  |  |
| <25 | 54.7 (108272) | 49.4 (303) | 54.8 (108328) | 34.7 (247) |
| 25-30 | 12.8 (25221) | 22.7 (139) | 12.7 (25160) | 28.1 (200) |
| 30-40 | 5.3 (10454) | 13.2 (81) | 5.3 (10401) | 18.8 (134) |
| >= 40 | 2.9 (5645) | 7.5 (46) | 2.8 (5596) | 13.3 (95) |
| Not recorded | 24.4 (48215) | 7.2 (44) | 24.4 (48223) | 5.1 (36) |
| **Smoking status** |  |  |  |  |
| Non smoker | 56.3 (111304) | 53.0 (325) | 56.2 (111189) | 61.8 (440) |
| Ex smoker | 17.5 (34649) | 41.6 (255) | 17.5 (34661) | 34.1 (243) |
| Light smoker | 9.0 (17712) | 3.1 (19) | 9.0 (17715) | 2.2 (16) |
| Moderate smoker | 1.7 (3266) | 1.0 (6) | 1.7 (3268) | <5 |
| Heavy smoker | 0.6 (1128) | <5 | 0.6 (1127) | <5 |
| Not recorded | 15.0 (29748) | 0.8 (5) | 15.0 (29748) | 0.7 (5) |
| **Geographical region** |  |  |  |  |
| East Midlands | 1.4 (2824) | 1.6 (10) | 1.4 (2826) | 1.1 (8) |
| East of England | 2.7 (5328) | 2.0 (12) | 2.7 (5328) | 1.7 (12) |
| London | 25.5 (50529) | 18.8 (115) | 25.5 (50510) | 18.8 (134) |
| North East | 3.1 (6220) | 6.5 (40) | 3.2 (6229) | 4.4 (31) |
| North West | 24.8 (49045) | 29.2 (179) | 24.8 (48990) | 32.9 (234) |
| South Central | 11.0 (21770) | 9.3 (57) | 11.0 (21738) | 12.5 (89) |
| South East | 10.9 (21478) | 12.4 (76) | 10.9 (21499) | 7.7 (55) |
| South West | 4.0 (7835) | 3.9 (24) | 4.0 (7832) | 3.8 (27) |
| West Midlands | 13.3 (26297) | 12.7 (78) | 13.3 (26270) | 14.7 (105) |
| Yorkshire & Humber | 3.3 (6481) | 3.6 (22) | 3.3 (6486) | 2.4 (17) |
| **Deprivation quintile** |  |  |  |  |
| 1 (least deprived) | 20.3 (40139) | 19.1 (117) | 20.3 (40121) | 18.9 (135) |
| 2 | 21.6 (42664) | 25.3 (155) | 21.6 (42667) | 21.4 (152) |
| 3 | 21.4 (42300) | 25.3 (155) | 21.4 (42277) | 25.0 (178) |
| 4 | 19.9 (39468) | 17.4 (107) | 19.9 (39449) | 17.7 (126) |
| 5 (most deprived) | 16.1 (31867) | 12.4 (76) | 16.1 (31826) | 16.4 (117) |
| Not recorded | 0.7 (1369) | <5 | 0.7 (1368) | <5 |
| **Comorbidities** |  |  |  |  |
| Asthma | 15.0 (29676) | 18.9 (116) | 15.0 (29645) | 20.6 (147) |
| COPD | 0.9 (1807) | 10.8 (66) | 0.9 (1840) | 4.6 (33) |
| Diabetes type 1 | 0.5 (1057) | <5 | 0.5 (1052) | 0.8 (6) |
| Diabetes type 2 | 4.6 (9088) | 32.8 (201) | 4.6 (9116) | 24.3 (173) |
| Hypertension | 9.8 (19295) | 55.6 (341) | 9.8 (19366) | 37.9 (270) |
| Parkinson | 0.1 (136) | 1.6 (10) | 0.1 (145) | <5 |
| Epilepsy | 1.1 (2136) | 2.3 (14) | 1.1 (2140) | 1.4 (10) |
| Cerebral palsy | 0.1 (180) | 0 | 0.1 (180) | 0 |
| Motor Neuron Disease | 0.0 (7) | <5 | 0.0 (8) | 0 |
| Huntington’s disease | 0.0 (6) | 0 | 0.0 (6) | 0 |
| Multiple sclerosis | 0.1 (265) | <5 | 0.1 (266) | <5 |
| Myasthenia | 0.0 (46) | <5 | 0.0 (47) | <5 |
| Downs syndrome | 0.0 (78) | <5 | 0.0 (80) | <5 |
| Learning disabilities excluding downs | 1.6 (3122) | 3.1 (19) | 1.6 (3127) | 2.0 (14) |

^*^SGTF status was used as a proxy for VOC B.1.1.7.

**Table S4: Demographic, medical characteristics and indicators of acute severity observed for critically ill patients tested positive in the community between 1^st^ November 2020 and 27^th^ January 2021 (critical care cohort) in the matched cohort dataset, by variant.**

|  | **Non-VOC B.1.1.7** | **VOC B.1.1.7^*^** | **Full Cohort** |
| --- | --- | --- | --- |
|  | *Cols% (counts)* | *Cols% (counts)* | *Cols% (counts)* |
| **Total number of patients** | 761 | 761 | 1522 |
| **Mean age (SD)** | 60.0 (12.6) | 57.6 (12.5) | 58.8 (12.6) |
| **Sex** |  |  |  |
| Female | 31.0 (236) | 36.7 (279) | 33.8 (515) |
| Male | 69.0 (525) | 63.3 (482) | 66.2 (1007) |
| **Ethnicity** |  |  |  |
| White | 64.7 (492) | 68.3 (520) | 66.5 (1012) |
| Indian | 5.8 (44) | 2.5 (19) | 4.1 (63) |
| Pakistani | 4.1 (31) | 6.4 (49) | 5.3 (80) |
| Bangladeshi | 2.5 (19) | 1.6 (12) | 2.0 (31) |
| Other Asian | 1.2 (9) | 3.2 (24) | 2.2 (33) |
| Caribbean | 0.0 (0) | 0.7 (5) | 0.3 (5) |
| Black African | 0.7 (5) | 1.6 (12) | 1.1 (17) |
| Chinese | 0.8 (6) | 0.1 (1) | 0.5 (7) |
| Other ethnic group | 4.1 (31) | 3.8 (29) | 3.9 (60) |
| Not recorded | 16.3 (124) | 11.8 (90) | 14.1 (214) |
| **Prior length of hospital stay** |  |  |  |
| Mean (SD) | 2.6 (5.0) | 2.3 (13.7) | 2.5 (10.3) |
| Median IQR | 1 (0 - 3) | 1 (0 - 3) | 1 (0 - 3) |
| **Dependency before admission to acute hospital care** |  |  |  |
| Able to live without assistance in daily activities | 70.8 (539) | 74.2 (565) | 72.5 (1104) |
| Some assistance in daily activities | 9.1 (69) | 6.6 (50) | 7.8 (119) |
| Total assistance with all daily activities | 0.9 (7) | <5 | 0.7 (10) |
| Not recorded | 19.2 (146) | 18.8 (143) | 19.0 (289) |
| **Severe comorbidities** |  |  |  |
| Cardiovascular | <5 | 1.1 (8) | 0.7 (10) |
| Respiratory | 2.5 (19) | 1.2 (9) | 1.8 (28) |
| Renal | 0.7 (5) | 0.8 (6) | 0.7 (11) |
| Liver | <5 | <5 | <5 |
| Metastatic disease | 1.3 (10) | <5 | 0.9 (13) |
| Haematological malignancy | 0.7 (5) | <5 | 0.5 (8) |
| Immunocompromised | 2.1 (16) | 1.1 (8) | 1.6 (24) |
| **BMI** |  |  |  |
| <25 | 38.1 (290) | 35.0 (266) | 36.5 (556) |
| 25 to <30 | 20.2 (154) | 21.9 (167) | 21.1 (321) |
| 30 to <40 | 30.0 (228) | 30.7 (234) | 30.4 (462) |
| 40+ | 11.7 (89) | 12.4 (94) | 12.0 (183) |
| **CPR within previous 21h** |  |  |  |
| In the community | 0.7 (5) | 0.3 (2) | 0.5 (7) |
| In the hospital | 1.3 (10) | 0.7 (5) | 1.0 (15) |
| No | 84.6 (644) | 86.6 (659) | 85.6 (1303) |
| Not recorded | 13.4 (102) | 12.5 (95) | 12.9 (197) |
| **Currently or recently pregnant** |  |  |  |
| Currently pregnant | <5 | <5 | 0.4 (6) |
| Recently pregnant (within 6 weeks) | <5 | 0.8 (6) | 0.5 (7) |
| Not known to be pregnant | 99.5 (757) | 98.8 (752) | 99.1 (1509) |
| **Invasively ventilated within first 24h** |  |  |  |
| No | 56.1 (427) | 58.6 (446) | 57.4 (873) |
| Yes | 22.2 (169) | 19.8 (151) | 21.0 (320) |
| Not recorded | 21.7 (165) | 21.6 (164) | 21.6 (329) |
| **APACHE II score** |  |  |  |
| Mean (SD) | 13.9 (5.0) | 12.7 (4.9) | 13.3 (5.0) |
| Median (IQR) | 13 (10 - 16) | 13 (10 - 16) | 13 (10 - 16) |
| **PaO2/FiO2 ratio** |  |  |  |
| Median (IQR) | 13.5 (9.5 – 18.3) | 13.0 (9.7 – 17.6) | 13.2 (9.7 – 18.0) |
| **PaO2/FiO2 ratio** |  |  |  |
| < 13.3 kPa (<100 mmHg) | 36.5 (278) | 38.2 (291) | 37.4 (569) |
| 13.3-16.6 kPa (100-200 mmHg) | 32.6 (248) | 32.6 (248) | 32.6 (496) |
| > 26.6 kPa (> 200 mmHg) | 5.5 (42) | 3.8 (29) | 4.7 (71) |
| Not recorded | 25.4 (193) | 25.4 (193) | 25.4 (386) |
| **FiO2** |  |  |  |
| Median (IQR) | 0.6 (0.5 – 0.75) | 0.6 (0.5 – 0.8) | 0.6 (0.50 – 0.75) |

^*^SGTF status was used as a proxy for VOC B.1.1.7

**Table S5: Demographic, medical characteristics and indicators of acute severity observed for critically ill patients tested positive in the community between 1^st^ November 2020 and 27^th^ January 2021 (critical care cohort) who have completed their critical care outcome (discharged alive or dead), by variant.**

|  | **Non-** **VOC B.1.1.7** | **VOC B.1.1.7^*^** | **Full Cohort** |
| --- | --- | --- | --- |
|  | *Cols% (counts)* | *Cols% (counts)* | *Cols% (counts)* |
| **Total number of patients** | 1235 | 1173 | 2408 |
| **Mean age (SD)** | 59.3 (12.9) | 57.3 (13.0) | 58.3 (13.0) |
| **Sex** |  |  |  |
| Female | 32.0 (395) | 33.9 (398) | 32.9 (793) |
| Male | 68.0 (840) | 66.1 (775) | 67.1 (1615) |
| **Ethnicity** |  |  |  |
| White | 72.0 (889) | 67.3 (790) | 69.7 (1679) |
| Indian | 4.2 (52) | 4.4 (52) | 4.3 (104) |
| Pakistani | 6.4 (79) | 4.3 (50) | 5.4 (129) |
| Bangladeshi | 1.8 (22) | 2.1 (25) | 2.0 (47) |
| Other Asian | 4.4 (54) | 4.3 (51) | 4.4 (105) |
| Caribbean | 0.8 (10) | 1.6 (19) | 1.2 (29) |
| Black African | 0.9 (11) | 1.7 (20) | 1.3 (31) |
| Chinese | 0.3 (4) | 0.5 (6) | 0.4 (10) |
| Other ethnic group | 4.1 (51) | 5.0 (59) | 4.6 (110) |
| Not recorded | 72.0 (889) | 67.3 (790) | 69.7 (1679) |
| **Prior length of hospital stay** |  |  |  |
| Mean (SD) | 2.2 (10.2) | 2.4 (14.7) | 2.3 (12.6) |
| Median IQR | 1 (0 - 3) | 1 (0 - 3) | 1 (0 - 3) |
| **Dependency before admission to acute hospital care** |  |  |  |
| Able to live without assistance in daily activities | 89.0 (1099) | 85.3 (1001) | 87.2 (2100) |
| Some assistance in daily activities | 8.0 (99) | 6.9 (81) | 7.5 (180) |
| Total assistance with all daily activities | 0.2 (2) | <5 | 0.2 (5) |
| Not recorded | 2.8 (381) | 7.5 (88) | 5.1 (123) |
| **Severe comorbidities** |  |  |  |
| Cardiovascular | <5 | 0.6 (7) | 0.4 (10) |
| Respiratory | 0.6 (8) | 0.9 (11) | 0.8 (19) |
| Renal | 0.6 (7) | <5 | 0.5 (11) |
| Liver | 0.5 (6) | <5 | 0.3 (7) |
| Metastatic disease | 0.4 (5) | <5 | 0.3 (7) |
| Haematological malignancy | 1.0 (12) | <5 | 0.6 (15) |
| Immunocompromised | 2.3 (28) | 1.2 (14) | 1.7 (42) |
| **BMI** |  |  |  |
| <25 | 22.2 (274) | 26.9 (315) | 24.5 (589) |
| 25 to <30 | 28.7 (355) | 24.6 (289) | 26.7 (644) |
| 30 to <40 | 36.6 (452) | 34.2 (401) | 35.4 (853) |
| 40+ | 12.5 (154) | 14.3 (168) | 13.4 (322) |
| **CPR within previous 21h** |  |  |  |
| In the community | 0.7 (9) | 0.5 (6) | 0.6 (15) |
| In the hospital | 0.6 (8) | 0.9 (11) | 0.8 (19) |
| No | 97.6 (1205) | 94.3 (1106) | 96.0 (2311) |
| Not recorded | 1.1 (13) | 4.2 (50) | 2.6 (63) |
| **Currently or recently pregnant** |  |  |  |
| Currently pregnant | 0.5 (6) | 0.5 (6) | 0.5 (12) |
| Recently pregnant (within 6 weeks) | 0.3 (4) | 0.6 (7) | 0.5 (11) |
| Not known to be pregnant | 99.2 (1225) | 98.9 (1160) | 99.0 (2385) |
| **Invasively ventilated within first 24h** |  |  |  |
| No | 76.1 (940) | 69.1 (811) | 72.7 (1751) |
| Yes | 20.5 (253) | 20.3 (238) | 20.4 (491) |
| Not recorded | 3.4 (42) | 10.5 (124) | 11.7 (166) |
| **APACHE II score** |  |  |  |
| Mean (SD) | 13.9 (5.0) | 12.7 (4.9) | 13.3 (5.0) |
| Median (IQR) | 14 (11 - 16) | 12 (10 - 15) | 13 (10 - 16) |
| **PaO2/FiO2 ratio** |  |  |  |
| Median (IQR) | 13.3 (10.0 – 18.2) | 13.5 (9.8 – 18.2) | 13.4 (10 – 18.2) |
| **PaO2/FiO2 ratio** |  |  |  |
| < 13.3 kPa (<100 mmHg) | 45.3 (559) | 40.2 (471) | 42.8 (1030) |
| 13.3-16.6 kPa (100-200 mmHg) | 38.8 (479) | 35.4 (415) | 37.1 (894) |
| > 26.6 kPa (> 200 mmHg) | 7.0 (87) | 6.6 (78) | 6.9 (165) |
| Not recorded | 8.9 (110) | 17.8 (209) | 13.2 (319) |
| **FiO2** |  |  |  |
| Median (IQR) | 0.6 (0.45 – 0.75) | 0.6 (0.4 – 0.76) | 0.6 (0.45 – 0.75) |

^*^SGTF status was used as a proxy for VOC B.1.1.7

**Table S6: Critical care outcomes observed for critically ill patients tested positive in the community between 1^st^ November 2020 and 27^th^ January 2021 (critical care cohort), by variant.**

|  | **Non-VOC B.1.1.7** | **VOC B.1.1.7^†^** | **Full Cohort** |
| --- | --- | --- | --- |
|  | *Cols% (counts)* | *Cols% (counts)* | *Cols% (counts)* |
| **Outcome at end of critical care** |  |  |  |
| Discharged | 56.5 (798) | 39.0 (788) | 46.2 (1586) |
| Died | 30.9 (437) | 19.1 (385) | 24.0 (822) |
| Still receiving critical care | 12.6 (178) | 41.9 (846) | 29.8 (1024) |
| **Duration of critical care***  **Days, median (IQR)** |  |  |  |
| Discharged patients | 6 (3 - 10) | 5 (3 - 8) | 5 (3 - 9) |
| Deaths | 13 (7 - 19) | 10 (5 - 15) | 10 (6 - 17) |
| **Organ support*** |  |  |  |
| No respiratory support | 2.3 (29) | 4.6 (54) | 3.4 (83) |
| Advanced respiratory support | 46.0 (568) | 42.7 (501) | 44.4 (1069) |
| Basic respiratory support | 86.2 (1064) | 81.8 (960) | 84.1 (2024) |
| No cardiovascular support | 5.5 (68) | 8.9 (104) | 7.1 (172) |
| Advanced cardiovascular support | 18.5 (229) | 18.7 (219) | 18.6 (448) |
| Basic cardiovascular support | 93.4 (1153) | 89.2 (1046) | 91.3 (2199) |
| Renal support | 13.5 (167) | 13.4 (157) | 13.5 (324) |
| Liver support | 2.6 (32) | 6.5 (76) | 4.5 (108) |
| Neurological support | 7.7 (95) | 9.2 (108) | 8.4 (203) |
| **Duration of organ support***  **Days, median (IQR)** |  |  |  |
| Advanced respiratory support | 11 (5 - 19) | 8 (5 - 13) | 9 (5 - 16) |
| Total (advanced + basic) respiratory support | 8 (4 - 15) | 6 (4 - 11) | 7 (4 - 13) |
| Advanced cardiovascular support | 2 (1 - 4) | 2 (1 - 4) | 2 (1 - 4) |
| Total (advanced + basic) cardiovascular support | 8 (5 - 15) | 6 (4 - 11) | 7 (4 - 13) |
| Renal support | 5.5 (3 - 10) | 4.5 (2 - 9) | 5 (2 - 10) |

* Among patients who have been discharged or died. **^†^**SGTF status was used as a proxy for VOC B.1.1.7

**Table S7: Demographics characteristics of primary care patients tested positive in hospital and community settings.**

|  | **Hospital tests** | **Community tests** |
| --- | --- | --- |
|  | *Cols% (counts)* | *Cols% (counts)* |
| **Total number of patients** | 15.9 (91965) | 85.1 (483667) |
| **Mean age (SD)** | 58.1 (23.4) | 38.1 (18.8) |
| **ICU admitted** | 4.3 (3998) | 0.3 (1538) |
| **SGTF** | 0.0 (16) | 24.8 (119863) |
| **Sex** |  |  |
| Female | 54.6 (50221) | 54.4 (263169) |
| Male | 45.4 (41744) | 45.6 (220498) |
| **Ethnicity** |  |  |
| White | 62.2 (57239) | 59.2 (286250) |
| Indian | 3.5 (3264) | 3.8 (18248) |
| Pakistani | 2.5 (2273) | 3.9 (18776) |
| Bangladeshi | 1.6 (1484) | 3.1 (14798) |
| Other Asian | 2.6 (2421) | 2.3 (10931) |
| Caribbean | 2.0 (1813) | 1.0 (4922) |
| Black African | 3.3 (3010) | 2.4 (11426) |
| Chinese | 0.3 (302) | 0.3 (1470) |
| Other ethnic group | 4.2 (3826) | 4.2 (20514) |
| Not recorded | 17.8 (16333) | 19.9 (96332) |
| **House size** |  |  |
| 1 person | 38.5 (35422) | 26.8 (129796) |
| 2 people | 22.9 (21048) | 19.8 (95754) |
| 3-5 people | 28.1 (25817) | 44.6 (215574) |
| 6-9 people | 4.5 (4120) | 6.7 (32212) |
| 10 or more | 6.0 (5558) | 2.1 (10331) |
| **House type** |  |  |
| Neither | 93.8 (86270) | 98.7 (477529) |
| Carehome | 5.9 (5457) | 1.2 (5725) |
| Homeless | 0.3 (238) | 0.1 (413) |
| **BMI** |  |  |
| <25 | 60.0 (55138) | 55.7 (269580) |
| 25-30 | 16.0 (14721) | 12.4 (59924) |
| 30-40 | 7.2 (6663) | 5.1 (24553) |
| >= 40 | 4.1 (3726) | 2.7 (13143) |
| Not recorded | 12.7 (11672) | 24.1 (116467) |
| **Smoking status** |  |  |
| Non smoker | 58.2 (53505) | 56.9 (275422) |
| Ex smoker | 27.0 (24828) | 17.1 (82925) |
| Light smoker | 7.3 (6728) | 9.1 (44237) |
| Moderate smoker | 1.4 (1282) | 1.6 (7905) |
| Heavy smoker | 0.8 (747) | 0.6 (2728) |
| Not recorded | 5.3 (4875) | 14.6 (70450) |
| **Geographical region** |  |  |
| East Midlands | 1.6 (1490) | 2.6 (12669) |
| East of England | 3.4 (3157) | 3.1 (15066) |
| London | 25.3 (23266) | 29.2 (141231) |
| North East | 3.0 (2772) | 2.4 (11678) |
| North West | 23.4 (21480) | 20.7 (100300) |
| South Central | 11.0 (10125) | 9.9 (47971) |
| South East | 11.2 (10267) | 11.7 (56709) |
| South West | 7.0 (6407) | 6.6 (31722) |
| West Midlands | 10.8 (9951) | 10.4 (50511) |
| Yorkshire & Humber | 3.3 (3050) | 3.3 (15810) |
| **Deprivation quintile** |  |  |
| 1 (least deprived) | 19.7 (18120) | 19.4 (93774) |
| 2 | 20.6 (18902) | 20.9 (101186) |
| 3 | 21.5 (19798) | 20.8 (100733) |
| 4 | 20.2 (18565) | 20.0 (96896) |
| 5 (most deprived) | 17.5 (16075) | 18.2 (87946) |
| Not recorded | 0.5 (505) | 0.6 (3132) |
| **Comorbidities** |  |  |
| Asthma | 15.2 (14011) | 14.7 (71129) |
| COPD | 7.0 (6463) | 1.1 (5115) |
| Diabetes type 1 | 0.9 (845) | 0.5 (2616) |
| Diabetes type 2 | 17.9 (16457) | 4.9 (23627) |
| Hypertension | 34.6 (31847) | 10.6 (51250) |
| Parkinson | 1.3 (1237) | 0.2 (793) |
| Epilepsy | 2.5 (2265) | 1.2 (5747) |
| Cerebral palsy | 0.2 (218) | 0.1 (538) |
| Motor Neuron Disease | 0.0 (41) | 0.0 (23) |
| Huntington’s disease | 0.0 (38) | 0.0 (53) |
| Multiple sclerosis | 0.4 (372) | 0.1 (720) |
| Myasthenia | 0.1 (93) | 0.0 (113) |
| Downs syndrome | 0.1 (122) | 0.1 (257) |
| Learning disabilities excluding Downs | 3.5 (3175) | 1.7 (8431) |

**Figure S1: (Top-left) Weekly positive tests by variant from 1^st^ November to 26^th^ January 2021, (top-right) weekly deaths by variant from 1^st^ November to 26^th^ January 2021, (bottom-left) weekly critical care admissions per variant from 1^st^ November to 26^th^ January 2021, (bottom-right) weekly deaths at the end of critical care by variant from 1^st^ November2020 to 27^th^ January 2021. SGTF status was used as a proxy for VOC B.1.1.7.**

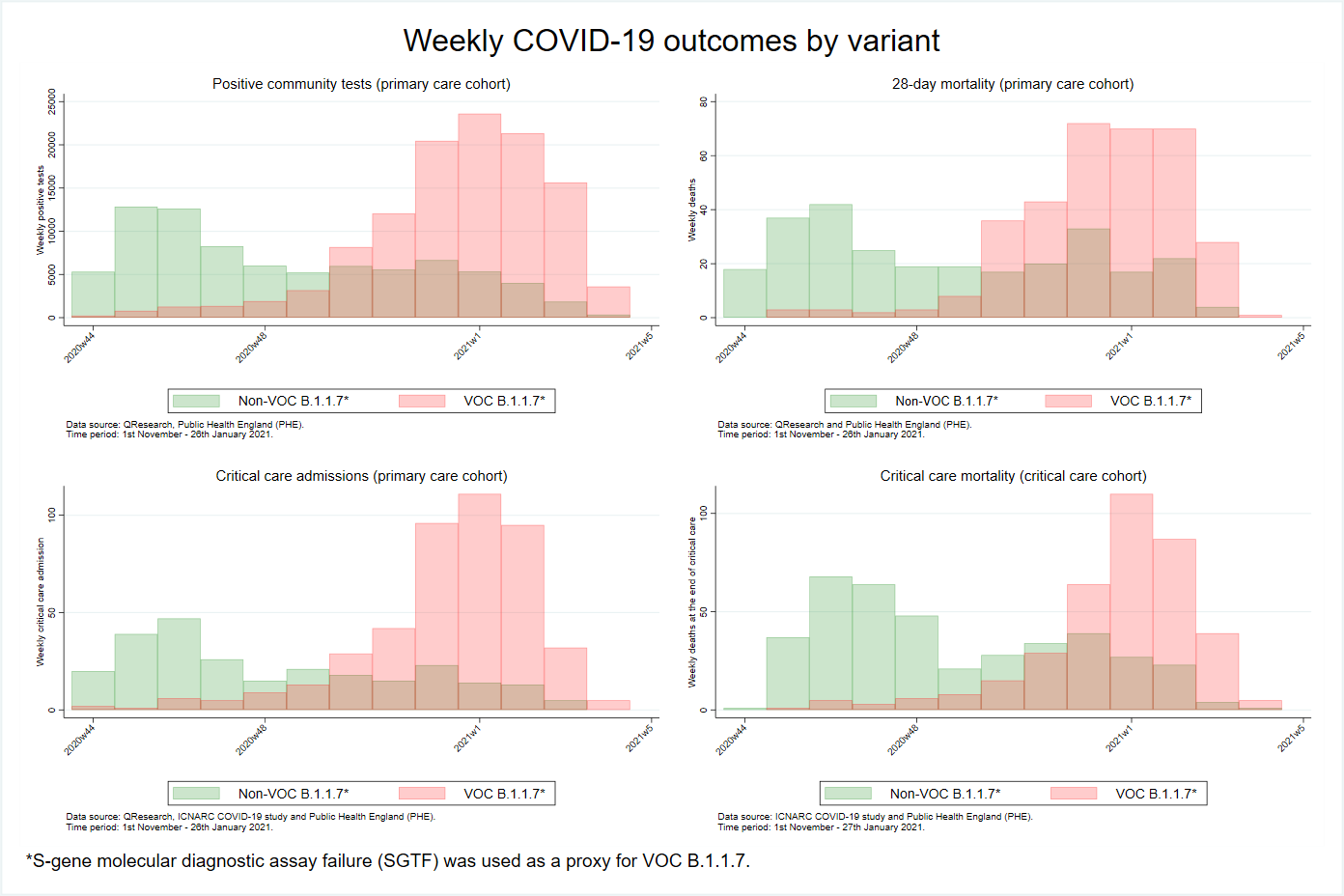
